# Evaluating causal associations of chronotype with pregnancy and perinatal outcomes and its interactions with insomnia and sleep duration: a Mendelian randomization study

**DOI:** 10.1101/2023.06.02.23290898

**Authors:** Qian Yang, Maria C Magnus, Fanny Kilpi, Gillian Santorelli, Ana Goncalves Soares, Jane West, Per Magnus, Siri E. Håberg, Kate Tilling, Deborah A Lawlor, M Carolina Borges, Eleanor Sanderson

## Abstract

**IMPORTANCE:** Observational studies suggest that chronotype is associated with pregnancy and perinatal outcomes. Whether these associations are causal is unclear.

**OBJECTIVE:** To explore associations of a lifetime genetic predisposition to an evening preference chronotype with pregnancy and perinatal outcomes, and explore differences in associations of insomnia and sleep duration with those outcomes between chronotype.

**DESIGN, SETTING, AND PARTICIPANTS:** We conducted two-sample Mendelian randomization (MR) using 105 genetic variants reported in a genome-wide association study (N=248 100) to instrument for lifelong predisposition to evening-versus morning-preference chronotypes. We generated variant-outcome associations in European ancestry women from UK Biobank (UKB, N=176 897), Avon Longitudinal Study of Parents and Children (ALSPAC, N=6826), Born in Bradford (BiB, N=2940) and Norwegian Mother, Father and Child Cohort Study (MoBa, with linked data from the Medical Birth Registry of Norway (MBRN), N=57 430), and extracted equivalent associations from FinnGen (N=190 879). We used inverse variance weighted (IVW) as main analysis, with weighted median and MR-Egger as sensitivity analyses. We also conducted IVW analyses of insomnia and sleep duration on the outcomes stratified by genetically predicted chronotype.

**EXPOSURES:** Self-reported and genetically predicted chronotype, insomnia and sleep duration.

**MAIN OUTCOMES AND MEASURES:** Stillbirth, miscarriage, preterm birth, gestational diabetes, hypertensive disorders of pregnancy, perinatal depression, low birthweight and macrosomia.

**RESULTS:** In IVW and sensitivity analyses we did not find robust evidence of effects of chronotype on the outcomes. Insomnia was associated with a higher risk of preterm birth among evening preference women (odds ratio 1.61, 95% confidence interval: 1.17, 2.21), but not among morning preference women (odds ratio 0.87, 95% confidence interval: 0.64, 1.18), with an interaction P-value=0.01. There was no evidence of interactions between insomnia and chronotype on other outcomes, or between sleep duration and chronotype on any outcomes.

**CONCLUSIONS AND RELEVANCE:** This study raises the possibility of a higher risk of preterm birth among women with insomnia who also have an evening preference chronotype. Our findings warrant replications due to imprecision of the estimates.

**Key points:** *Question:* Does an evening preference chronotype adversely affect pregnancy and perinatal outcomes? Is there an interaction between chronotype and either insomnia or sleep duration in relation to those outcomes?

*Findings:* There was no evidence that evening preference was associated with pregnancy or perinatal outcomes. Women with a genetically predicted insomnia had a higher risk of preterm birth, if they also had a genetically predicted preference for evening chronotype.

*Meaning:* The suggestive interaction between insomnia and evening preference on preterm birth, if replicated, supports targeting insomnia prevention in women of reproductive age with an evening chronotype.

## Introduction

Adverse pregnancy and perinatal health conditions, e.g. stillbirth, miscarriage, preterm birth (PTB), gestational diabetes, hypertensive disorders of pregnancy (HDP), perinatal depression, and small-/large-for-gestational age, affect up to 40% of pregnancies.^1, 2^ Sleep traits, in particular insomnia and short/long sleep duration, have been found to associate with many of these outcomes in systematic reviews of observational studies using traditional multivariable regression analyses.^3, 4^ Mendelian randomization (MR) is less prone to confounding than observational studies, as genetic variants which are randomly allocated at meiosis are used as instrumental variables (IVs).^5^ Recent MR studies support associations of insomnia with miscarriage, perinatal depression and low offspring birthweight (LBW),^6^ shorter sleep duration with perinatal depression, and shorter and longer sleep duration with LBW.^7^

Chronotype refers to a person’s circadian preference, defined as morning-, evening- or no-preference. Chronotype is assessed either by self-report or actigraphy-defined timing of when the person is most active.^8–14^ Observational studies suggest that late sleep midpoint (reflecting evening preference) is associated with an increased risk of gestational diabetes in a US cohort (N=7524) and its subsample of actigraphy data (N=782), but not HDP in the same participants.^8, 9^ Late sleep midpoint was not associated with pregnancy loss in another US cohort (N=1088),^10^ or small-for-gestational age in a Chinese cohort (N=11 192).^11^ The first US cohort also showed an increased risk of PTB in women with late sleep midpoints (>5 a.m.),^12^ while a Chinese cohort showed the opposite, with an increased risk of PTB among those with early midpoints (<2.45 a.m.).^11^ Studies of perinatal depression are inconclusive due to small sample sizes (N=51 and 179).^13, 14^ To the best of our knowledge, we have found no MR studies of chronotype on pregnancy/perinatal outcomes.^15^

Poor sleep quality and unhealthy sleep duration have been observed within certain groups of chronotype preferences in previous studies of pregnant people,^9, 10, 13, 14^ and large studies of non-pregnant people.^16, 17^ For example, participants whose preference is evening but have to be active early morning, may have poorer quality and shorter sleep duration. Examining interactions of insomnia and sleep duration with chronotype could also reveal novel associations not seen when these sleep traits are studied independently of chronotype.

The aims of this study were to 1) explore associations of a lifetime genetic predisposition to evening-versus morning-preference on stillbirth, miscarriage, gestational diabetes, HDP, perinatal depression, PTB, LBW, macrosomia, and birthweight using two-sample MR; and 2) explore differences in associations of insomnia and sleep duration on these outcomes between women with morning- and evening-preference.

## Methods

### Participants

This study used data from the MR-PREG collaboration, which aims to explore causes and consequences of different pregnancy and perinatal outcomes.^6, 7, 18^ We used individual-level data from UK Biobank (UKB, N=176 897 women who had experienced at least one pregnancy), and mother-offspring pairs from Avon Longitudinal Study of Parents and Children (ALSPAC, N=6826), Born in Bradford (BiB, N=2940) and Norwegian Mother, Father and Child Cohort Study (MoBa, N=57 430), and summary-level data from FinnGen (the nationwide network of Finnish biobank with 173 746 women). FinnGen is a large repository of summary gene-disease association data and for pregnancy conditions the control groups include women who have never been pregnant.^19^ Figure 1 shows how each cohort contributed to our MR analyses. All studies had ethical approval from relevant national or local bodies and participants provided written informed consent. Details of their recruitment and genotyping are described in eAppendix 1 in Supplement.

**Figure 1.**
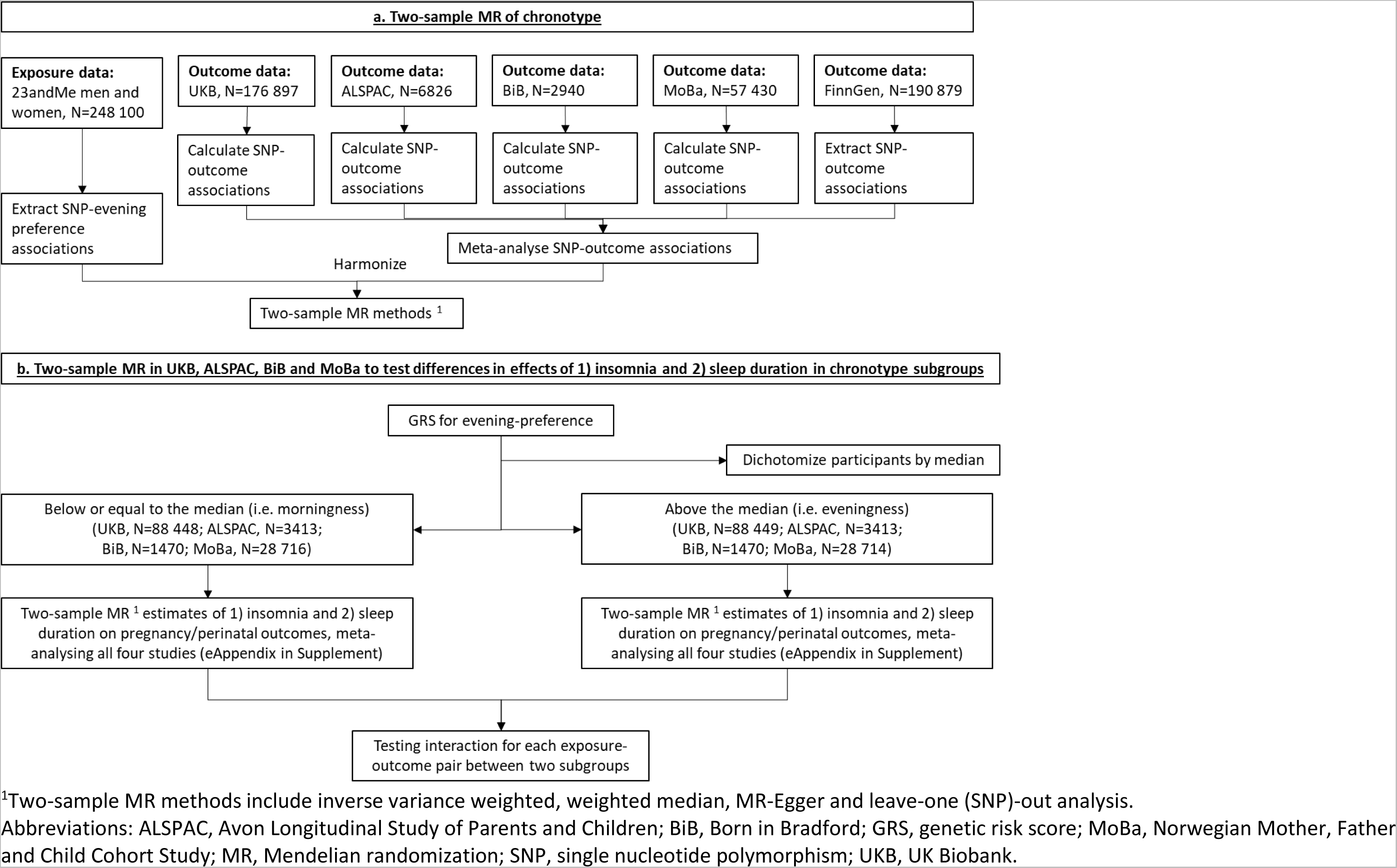
Summary of methods and data contributing to our two-sample MR analyses

### Pregnancy and perinatal outcomes

We included ever experiencing stillbirth, ever experiencing miscarriage, and PTB (gestational age <37 completed weeks), gestational diabetes, HDP, perinatal depression, LBW (birthweight <2500 grams), macrosomia (birthweight >4500 grams), and birthweight as a continuous measure. Full details about how these outcomes were measured and derived in each participating study and how we harmonised them across studies are in eTable 1 in Supplement. In UKB, gestational age was only available for a subset of women (N=5362) who had a child born during or after 1989, the earliest date for which linked electronic hospital perinatal data were available. Therefore, we *a priori* decided to examine associations with LBW and macrosomia rather than small- and large-for-gestational age. If multiple pregnancies were enrolled in the birth cohorts, we randomly selected one pregnancy per woman. In FinnGen, it was only possible to study miscarriage, gestational diabetes, HDP and PTB, where women with pre-existing hypertension were included in HDP cases.

### Chronotype and IVs

The most recent genome-wide association study (GWAS) of chronotype combines data from UKB and 23andMe, though sex-specific results were not available.^20^ We extracted summary results of genome-wide significant associations available in 23andMe (without UKB) participants for two-sample MR, to minimize potential bias due to its overlap with our outcome sample.^21^ Among the 248 100 23andMe participants included in the most recent GWAS, ≥97% of them were of European ancestry, and ∼44.8% of them were female.

In 23andMe, the question used to measure chronotype (“Are you naturally a night person or a morning person?”) was asked in two surveys.^20, 22^ Response options were initially “night owl”, “early bird”, and “neither”, but changed to “night person”, “morning person”, “neither”, “it depends”, and “I’m not sure”.^20, 22^ As shown in eTable 2 in Supplement, participants with discordant (morning preference in one survey but evening preference in the other) or neutral (“neither”, “it depends”, and “I’m not sure”) answers to both questions were excluded from the GWAS,^20^ who accounted for ∼13% of 23andMe participants.^22^ Participants with a neutral and a non-neutral (“night owl”, “early bird”, “night person” and “morning person”) answers were included solely based on the non-neutral one.^20^ Thus, the GWAS included 127 622 and 120 478 individuals of evening and morning preference, respectively, and identified 110 genome-wide significant (P-value<5×10^-8^) single nucleotide polymorphisms (SNPs) in autosomes using linear mixed model.^20^ We used the ‘clumping’ function from ‘TwoSampleMR’ R package to check if those SNPs were independent,^23^ and 105 SNPs (eTable 3 in Supplement) were retained based on a threshold of R^2^<0.01 using all European samples from the 1000 Genomes Project as the reference population.

### Insomnia, sleep duration and IVs

Information on insomnia and sleep duration was collected at the UKB initial assessment centre, and described in eAppendix 1 in Supplement. Characteristics of genome-wide significant SNPs for insomnia (81 SNPs) and sleep duration (78 SNPs) were extracted from the largest GWAS available,^24, 25^ and listed in eTable 4 in Supplementary. None of these SNPs were overlapped with the 105 SNPs for chronotype. Insomnia (r_g_=-0.10, P-value=0.113) and sleep duration (r_g_=0.11, P-value=0.142) were genetically weakly correlated with chronotype in UKB women using linkage disequilibrium score regression,^26^ and their IVs have been used in previous MR studies.^6, 7^

### Statistical analyses

#### Two-sample MR of chronotype

The 105 SNP-chronotype associations were from the GWAS in 23andMe.^20^ We generated SNP-outcome associations in UKB, ALSPAC, BiB and MoBa using logistic regression for binary outcomes and linear regression for birthweight, adjusting for genotyping batch (only applicable in UKB and MoBa), women’s age and top 10 principal components. We also extracted female-specific associations of the 105 SNPs with miscarriage, gestational diabetes, HDP and PTB from summary-level data of FinnGen,^19^ and meta-analysed SNP-outcome associations across five studies using fixed-effects with inverse variance weights.

We used MR inverse variance weighted (IVW) method as main analysis, which meta-analyses each coefficient calculated as the SNP-outcome association divided by the SNP-exposure association with a fixed effect model.^27^ We repeated IVW analyses by leaving one cohort out in turn to explore between-study heterogeneity. To explore the strength of IVs, we calculated the proportion of variance of evening-preference and related mean F-statistic of the 105 SNPs.^28^ We further compared the sex-combined associations from 23andMe with the equivalent associations in UKB women, whose measurement of chronotype was described in eAppendix 1 in Supplement. To explore potential unbalanced horizontal pleiotropy, sensitivity analyses included estimations of between-SNP heterogeneity using Q-statistic and leave-one (SNP)-out analysis, and MR with weighted median and MR-Egger approaches.^29, 30^ The association of maternal with fetal genotype for sleep traits might also introduce bias for any outcomes in the index pregnancy, which are plausibly influenced by fetal genotype.^31^ To explore this, we used data from MoBa, where trio data were available, to compare SNP-outcome (except for stillbirth and miscarriage history) associations with adjustments for (1) fetal genotypes (N=43 649), (2) both fetal and paternal genotypes (N=28 214), versus (3) without those adjustments (N=57 430).

#### Two-sample MR stratified on genetically predicted chronotype

We compared associations of insomnia and sleep duration with the outcomes between women with a genetically predicted evening-versus morning-preference (Figure 1). The analyses were undertaken on individual-level data in UKB and birth cohorts. We constructed a weighted genetic risk score of the 105 genome-wide significant SNPs for evening preference using external weights from 23andMe, and separate women into two subgroups using its median (eFigure 1 in Supplementary).^32^

Among women below and above the median, we obtained two-sample MR IVW estimates of insomnia and sleep duration for each outcome (eFigure 2 in Supplementary). Briefly, in UKB we generated SNP-exposure and SNP-outcome associations in a split cross-over samples design.^33^ We also conducted two-sample MR using SNP-exposure associations from UKB, and SNP-outcome associations from birth cohorts, and further meta-analysed MR estimates from all four cohorts using fixed-effects with inverse variance weights (eAppendix 2 in Supplement).^6, 7^ Differences between results among women below and above the median were calculated using z-score of the ratio of odds ratios (ORs, i.e. 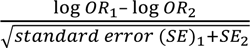, referring to the normal distribution).^34^ For birthweight, the same approach was taken using the difference in differences in mean between the two subgroups.^34^

Two-sample MR analyses were conducted using ‘TwoSampleMR’ R package.^23^ All analyses were performed using R 3.6.1 (R Foundation for Statistical Computing, Vienna, Austria).

## Results

Table 1 summarizes characteristics of included women from UKB, ALSPAC, BiB, MoBa and FinnGen. In two-sample MR, the 105 SNPs explained approximately 0.68% of the variance in chronotype (eTable 3 in Supplement), and their mean F-statistic was 16.1 among 248 100 participants. The association of all of these 105 SNPs with evening preference in 176 887 UKB women was directionally consistent, though weaker, than the equivalent using GWAS 23andMe women and men (eFigure 3 in Supplement).

**Table 1.** Characteristics of women from UKB, ALSPAC, BiB, MoBa and FinnGen included in this study

| Variable | UKB (N=176 897) | ALSPAC (N=6826) | BiB (N=2940) | MoBa (N=57 430) | FinnGen (N≤190 879) |
| --- | --- | --- | --- | --- | --- |
| Country | UK | UK | UK | Norway | Finland |
| Year of recruitment | 2006-2010 | 1991-1992 | 2007-2010 | 1999-2009 | 1986-present |
|  | *Mean (standard deviation)* |  |  |  |  |
| Age at delivery <sup>1</sup> | 25.5 (4.6) years | 28.7 (4.7) years | 26.8 (6.0) years | 30.4 (15.1) years | Not available |
| Gestational age | 38.9 (3.8) weeks | 39.6 (1.7) weeks | 39.7 (1.9) weeks | 39.4 (2.1) weeks | Not available |
| Offspring birthweight | 3186.7 (547.6) grams | 3441.5 (523.0) grams | 3357.9 (571.2) grams | 3600.0 (562.1) grams | Not available |
| Sleep duration | 7.2 (1.1) hours | Not available | Not available | 8.0 (1.5) hours <sup>2</sup> | Not available |
|  | *N (%)* |  |  |  |  |
| Offspring sex, male | Not available | 3430 (50.2) | 1504 (51.2) | 29 315 (51.0) | Not available |
| Having morning preference | 103 190 (58.3) | Not available | Not available | Not available | Not available |
| Having insomnia | 57 318 (32.4) | 909 (13.3) <sup>2</sup> | Not available | Not available | Not available |
|  | *N cases/N controls (prevalence, %)* |  |  |  |  |
| Ever experiencing stillbirth | 4907/107 791 (4.4) | 48/4546 (1.0) | 31/2588 (1.2) | 255/39 560 (0.6) | Not available |
| Ever experiencing miscarriage | 42 717/107 791 (28.4) | 1378/4546 (23.3) | 14/2588 (0.5) | 10 090/39 560 (20.3) | 15 073/135 962 (10.0) |
| Preterm birth | 551/4811 (10.3) | 285/4931 (5.5) | 172/2706 (6.0) | 2879/49 586 (5.5) | 8108/135 806 (5.6) |
| Gestational diabetes | 726/170 308 (0.4) | 34/6283 (0.5) | 136/2657 (4.9) | 470/56 524 (0.8) | 11 279/179 600 (5.9) |
| HDP | 2128/174 769 (1.2) | 1099/5698 (16.2) | 347/2159 (13.8) | 7300/49 866 (12.8) | 13 071/177 808 (6.8) |
| Perinatal depression | 5168/20 860 (19.9) | 423/5896 (6.2) | 312/2245 (12.2) | 2554/49 977 (4.9) | Not available |
| Low offspring birthweight | 13 429/149 084 (8.3) | 337/6376 (5.0) | 167/2725 (5.8) | 1675/52 905 (3.1) | Not available |
| Macrosomia | 2716/149 084 (1.8) | 113/6376 (1.7) | 42/2725 (1.5) | 2468/52 905 (4.5) | Not available |

<sup>1</sup> UKB women were recruited with an average age 56.9 (standard deviation 7.8) years, and FinnGen participants were recruited with an median age of 63 years.<sup>19</sup>
<sup>2</sup> Data were not used in this two-sample MR.
Abbreviations: ALSPAC, Avon Longitudinal Study of Parents and Children; BiB, Born in Bradford; GRS, genetic risk score; HDP, hypertensive disorders of pregnancy; MoBa, the Norwegian Mother, Father and Child Cohort; UKB, UK Biobank.

In the main IVW analyses combining the five studies, there was little evidence of associations of chronotype with the outcomes with all 95% confidence intervals (CIs) including the null (Figure 2). There was no strong statistical evidence of between-study heterogeneity, and the removal of the largest studies (UKB or FinnGen) did not materially change the null effect, though resulted in wider CIs, as would be expected (eFigure 4 in Supplement). Between-SNP heterogeneity was observed for miscarriage, gestational diabetes, HDP, LBW and birthweight (Figure 2), as a few individual SNPs showed associations with the same outcomes (eFigure 5 in Supplement). However, leave-one (SNP)-out analyses suggested little evidence of a single SNP driving our IVW results (eFigure 6 in Supplement). Furthermore, sensitivity analyses using weighted median and MR-Egger approaches were consistent with the null associations for all outcomes, and the MR-Egger intercept did not provide evidence of unbalanced horizontal pleiotropy for any outcomes (Figure 2). After adjusting for fetal genotype or both fetal and paternal genotype in MoBa, most SNP-outcome associations were statistically consistent with the unadjusted results (i.e. 95% CI for the difference between two associations including the null), with exceptions for ≤2 SNPs with gestational diabetes, HDP, perinatal depression, LBW, macrosomia and birthweight (eFigure 7 and eTable 5 in Supplement).

**Figure 2.**
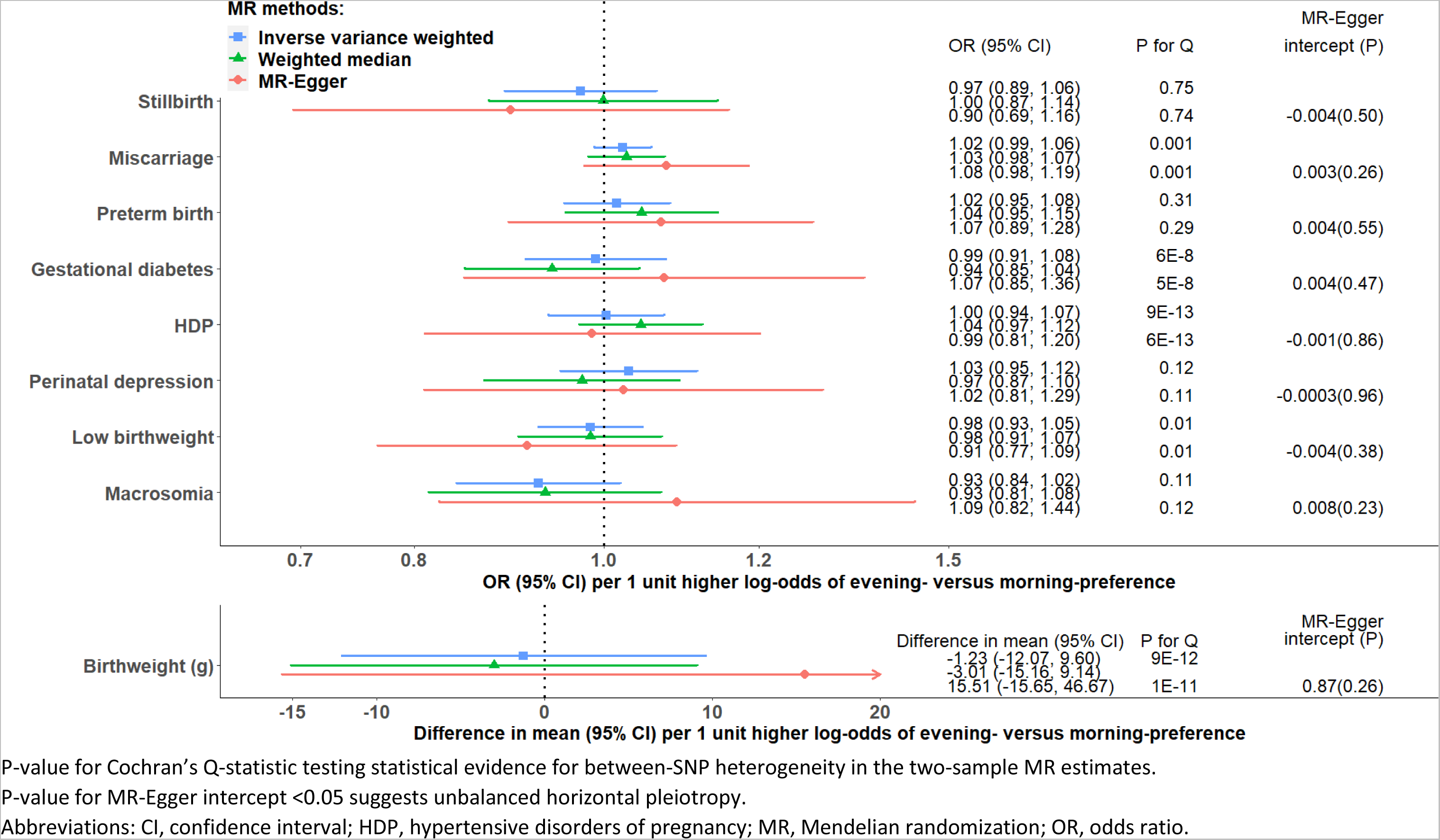
Two-sample MR estimates for causal effects of chronotype on pregnancy and perinatal outcomes meta-analysing UK Biobank, three birth cohorts and FinnGen

After combining all individual-level data (i.e. UKB, ALSPAC, BiB and MoBa), insomnia was associated with PTB among women with genetic predisposition to evening preference (OR 1.61, 95% CI: 1.17, 2.21), but not among morning preference women (OR 0.87, 95% CI: 0.64, 1.18). We observed a significant interaction between these two sleep traits on risk of PTB (P-value=0.02). Associations of insomnia with other outcomes (Figure 3) and sleep duration with all outcomes (Figure 4) were similar in morning-versus evening-preference subgroups (interaction P-value>0.05).

**Figure 3.**
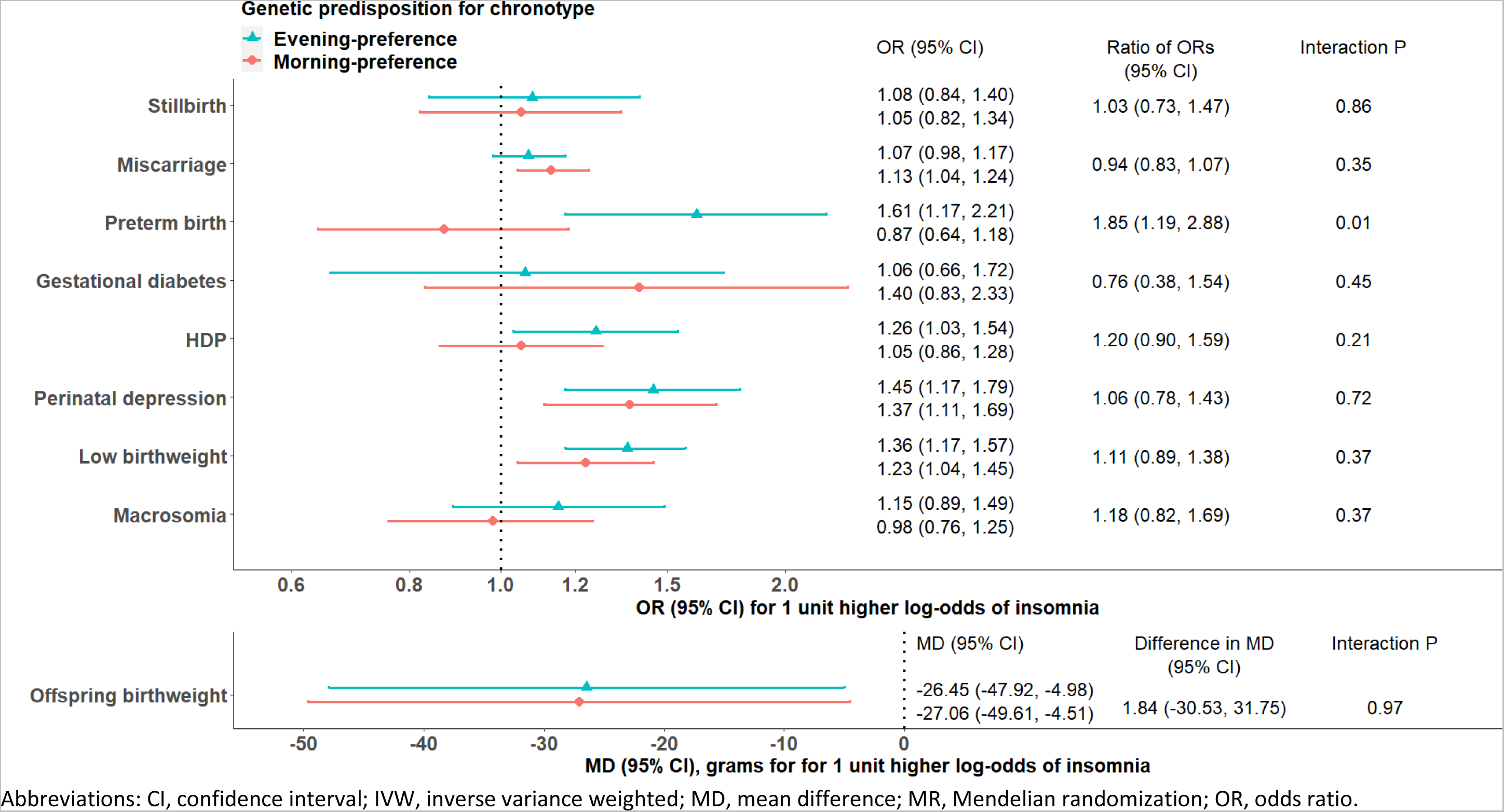
Two-sample MR IVW estimates meta-analysing UK Biobank and three birth cohorts for causal effects of insomnia on pregnancy and perinatal outcomes, stratified on chronotype

**Figure 4.**
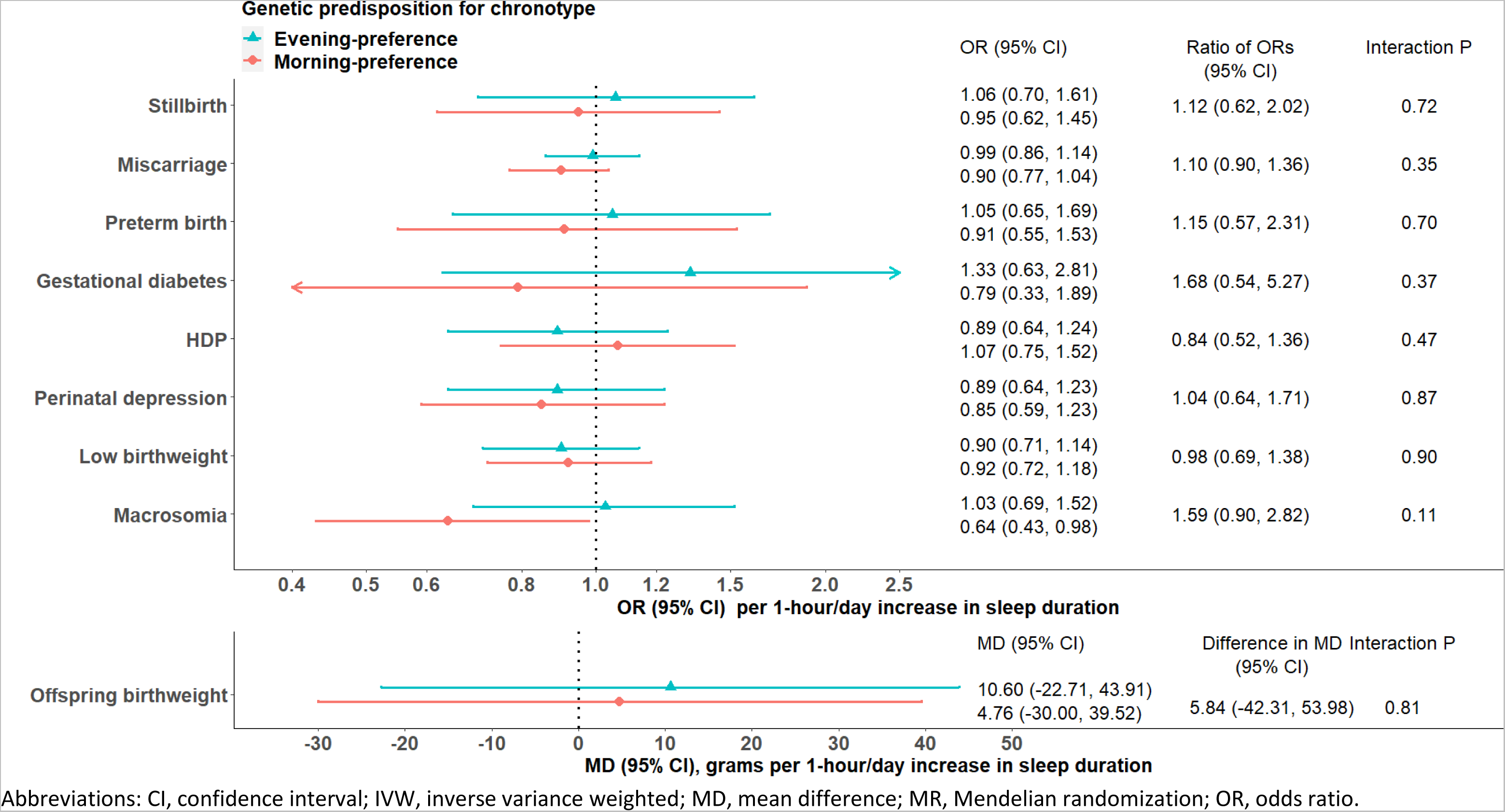
Two-sample MR IVW estimates meta-analysing UK Biobank and three birth cohorts for causal effects of sleep duration on pregnancy and perinatal outcomes, stratified on chronotype

## Discussion

Our study used a novel application of MR to explore associations of maternal chronotype with adverse pregnancy and perinatal outcomes, and whether associations of insomnia and sleep duration with these outcomes differ between chronotype preferences. We interpret our results as estimating the association of a lifetime genetic predisposition to a trait (e.g. evening-(versus morning-) preference) on the basis that SNPs are determined at conception.^35, 36^ In comparison to other sleep related traits, few studies have explored associations of chronotype with pregnancy and perinatal outcomes, with details summarized in eTable 6 in Supplement. Thus our results (particularly the suggestive interaction between insomnia and chronotype for PTB) need further replications.

Our null associations of chronotype with stillbirth history, miscarriage history, HDP and LBW were broadly consistent with previous observational studies of pregnancy loss,^10^ HDP,^8, 9^ and small-for-gestational age.^11^ Our null association of chronotype with perinatal depression was largely in agreement with two small observational studies of self-report chronotype on depressive symptoms scores from 2^nd^ trimester to 16 weeks postpartum.^13, 14^ One exception was at 2 weeks postpartum, when a statistically significant higher score (indicating severer depressive symptoms) was observed among women with evening-(versus morning-) preference.^14^ In contrast to our findings, a US prospective cohort study found self-report evening preference was associated with higher risks of gestational diabetes and PTB,^8, 12^ though a Chinese cohort found that morning preference, assessed using accelerometer, was associated with a higher risk of PTB.^11^ Differences between these two cohorts, and between them and our MR results could reflect differences in how chronotype was measured, residual confounding, and true population differences. They could also reflect the fact that the observational studies explored associations of chronotype assessed in pregnancy, where as our MR analyses are of genetically predicted lifetime exposure to chronotype possibility with potential horizontal pleiotropy (discussed in limitations).

Emerging evidence suggests that when people’s work and childcare requirements contradict their chronotype preference (a phenomenon known as ‘social jetlag’), there can be adverse consequences on cardiometabolic health,^37–40^ which are strongly correlated to gestational diabetes and HDP in women.^41, 42^ We only identified three small birth cohorts investigating associations between social jetlag and our outcomes, which was consistently defined as a difference in sleep midpoints (median times of self-report sleep and wake up times on a 24-hour clock) between free-day and work-day.^10, 13, 43^ These studies found little evidence for associations of social jetlag with pregnancy loss or perinatal depression,^10, 13^ but a higher risk of gestational diabetes with inflated 95% CIs in a subgroup of ≥2-hour social jetlag.^43^

Given social jetlag could be correlated with sleep debt and poor sleep quality,^40, 44^ we used two-sample MR stratified on genetically predicted chronotype in individual-level data to test chronotype-by-insomnia and -by-sleep duration interactions. Consistent with our previous MR study,^6^ insomnia was associated with higher risks of miscarriage, perinatal depression, and LBW in both morning- and evening-preference subgroups with little evidence for interactions. We identified an interaction between chronotype and insomnia with PTB, while our previous MR study found little evidence that insomnia was associated with PTB.^6^ The interaction we observed might provide an approach of identifying women at risk of PTB early in pregnancy, by monitoring women with both evening preference and insomnia. Randomized controlled trials showed that cognitive behavioural therapy for insomnia can effectively reduce insomnia in pregnancy, but they explored its effect on perinatal depression only.^45–47^ Our findings might support those trials to further explore its effects on PTB particularly in women with evening preference.

### Strengths and limitations

The main strengths of our study included 1) an exploration of multiple pregnancy and perinatal outcomes of clinical importance and public health concern; 2) use of MR to explore associations of a lifetime predisposition to morning or evening preference; 3) exploring whether associations of insomnia and sleep duration with those outcomes interact with chronotype; 4) a number of sensitivity analyses to explore how robust our results were to MR assumptions; and 5) large sample sizes for some important outcomes, e.g. ever having a miscarriage, which have not been explored previously in observational or MR studies.

Our two-sample MR may be vulnerable to weak instrument bias,^21^ and we were not able to test the relevance of genetic IVs for chronotype in pregnant people as none of our birth cohorts collected this information. We showed directionally consistent, though weaker, associations with chronotype in UKB women versus the combined 23andMe women and men. As we used SNPs selected from 23andMe women and men as IVs, if there was a true weaker IV-exposure (chronotype) effect in women, we may have underestimate potential effects of lifetime evening preference on pregnancy and perinatal outcomes. Our results may be biased by unbalanced horizontal pleiotropy, as some SNPs that we used as instrumental variables for chronotype are known to genome-wide significantly associate with education attainment, anthropometric measures, and other health related factors.^48^ However, none of our sensitivity analyses provided evidence of substantial bias due to unbalanced horizontal pleiotropy. A monotonicity assumption is required for our MR estimates to be interpreted as a local average treatment effects.^5^ This means our genetic IVs for sleep traits (e.g. chronotype) cannot increase the probability of reported exposure (e.g. evening preference) in some women while decrease it in others,^49, 50^ though it is difficult to evaluate the influence of violating this assumption on MR studies.

We did not apply any corrections for multiple testing, as it is more relevant to a hypothesis-searching study (e.g. GWAS) than to our hypothesis-driven design.^35^ We acknowledge that the interaction we observed could be due to chance, and emphasized the importance of further replications with sufficient cases to secure statistical power, and triangulation of evidence across different methods with different sources of bias.^51^ Measurement errors in our self-report sleep traits, and potential selection bias in UKB might bias MR results, which have been comprehensively discussed in our previous MR studies.^6, 7^

## Conclusions

Our findings suggest little evidence that a genetic lifetime predisposition to evening compared to morning preference was associated with pregnancy and perinatal outcomes, but raise the possibility that women with predispositions to an evening preference and insomnia are at a higher risk of PTB. These findings require replications due to their uncertainty as reflected by wide CIs for some outcomes, and exploration in non-European populations.

## Data Availability Statement

We used both individual participant cohort data and publicly available summary statistics. We present summary statistics that we generated from those individual participant cohort data in eTables 7-10 in Supplement. Full information on how to access UKB data can be found at its website (https://www.ukbiobank.ac.uk/researchers/). All ALSPAC data are available to scientists on request to the ALSPAC Executive via this website (http://www.bristol.ac.uk/alspac/researchers/), which also provides full details and distributions of the ALSPAC study variables. Similarly, data from BiB are available on request to the BiB Executive (https://borninbradford.nhs.uk/research/how-to-access-data/). Data from MoBa are available from the Norwegian Institute of Public Health after application to the MoBa Scientific Management Group (see its website https://www.fhi.no/en/op/data-access-from-health-registries-health-studies-and-biobanks/data-access/applying-for-access-to-data/ for details). Summary statistics from FinnGen are publicly available on its website (https://finngen.gitbook.io/documentation/data-download).

## Funding

This work was supported by the University of Bristol and UK Medical Research Council (MM_UU_00011/1, MM_UU_00011/3 and MM_UU_00011/6), the US National Institute for Health (R01 DK10324), the European Research Council via Advanced Grant 669545, the British Heart Foundation (AA/18/7/34219 and CS/16/4/32482) and the National Institute of Health Research Bristol Biomedical Research Centre at University Hospitals Bristol NHS Foundation Trust and the University of Bristol. Q.Y. is funded by a China Scholarship Council PhD Scholarship (CSC201808060273). M.C.B. was funded by a UK Medical Research Council Skills Development Fellowship (MR/P014054/1) and a University of Bristol Vice Chancellor Fellowship during her contribution to this research. M.C.M. has received funding from the European Research Council under the European Union’s Horizon 2020 research and innovation programme (grant agreement No 947684). M.C.M and S.E.H are partly funded by the Research Council of Norway through its Centres of Excellence funding scheme (project No 262700) and by the Research Council of Norway (project no. 320656) and co-funded by the European Union (ERC, BIOSFER, 101071773). D.A.L. is a British Heart Foundation Chair (CH/F/20/90003) and a National Institute of Health Research Senior Investigator (NF-0616-10102).

The UK Medical Research Council and Wellcome (Grant ref: 217065/Z/19/Z) and the University of Bristol provide core support for ALSPAC. This publication is the work of the authors and D.A.L will serve as guarantor for the contents of this paper. A comprehensive list of grants funding is available on the ALSPAC website (http://www.bristol.ac.uk/alspac/external/documents/grant-acknowledgements.pdf); This research was specifically funded by Wellcome Trust (WT088806), and child’s GWAS data was generated by Sample Logistics and Genotyping Facilities at Wellcome Sanger Institute and LabCorp (Laboratory Corporation of America) using support from 23andMe. BiB receives core funding from the Wellcome Trust (WT101597MA), a joint grant from the UK Medical and Economic and Social Science Research Councils (MR/N024397/1), British Heart Foundation (CS/16/4/32482), and the National Institute of Health Research under its Applied Research Collaboration for Yorkshire and Humber and Clinical Research Network research delivery support. Further support for genome-wide and multiple ‘omics measurements in BiB is from the UK Medical Research Council (G0600705), National Institute of Health Research (NF-SI-0611-10196), US National Institute of Health (R01DK10324), and the European Research Council under the European Union’s Seventh Framework Programme (FP7/2007–2013) / ERC grant agreement no 669545.

The funders had no role in the design of the study; the collection, analysis, or interpretation of the data; the writing of the manuscript; or the decision to submit the manuscript for publication. The views expressed in this paper are those of the authors and not necessarily those of any funder. Neither the European Union nor the granting authority can be held responsible for them.

## Competing interests

KT has acted as a consultant for CHDI Foundation, and Expert Witness to the High Court in England, called by the UK Medicines and Healthcare products Regulatory Agency, defendants in a case on hormonal pregnancy tests and congenital anomalies 2021/22. DAL has received support from Medtronic LTD and Roche Diagnostics for biomarker research that is not related to the study presented in this paper. The other authors report no conflicts.

## Acknowledgments

This research has been conducted using the UKB Resources under application number 23938. The authors would like to thank the participants and researchers from UKB who contributed or collected data. We are extremely grateful to all the families who took part in this study, the midwives for their help in recruiting them, and the whole ALSPAC team, which includes interviewers, computer and laboratory technicians, clerical workers, research scientists, volunteers, managers, receptionists and nurses. BiB is only possible because of the enthusiasm and commitment of the children and parents in BiB. We are grateful to all the participants, health professionals, schools and researchers who have made BiB happen. This research has been conducted using MoBa data using application number 2552. MoBa is supported by the Norwegian Ministry of Health and Care services and the Ministry of Education and Research. We are grateful to all the participating families in Norway who take part in this on-going cohort study. We thank the Norwegian Institute of Public Health (NIPH) for generating high-quality genomic data. This research is part of the HARVEST collaboration, supported by the Research Council of Norway (#229624). We also thank the NORMENT Centre for providing genotype data, funded by the Research Council of Norway (#223273), South East Norway Health Authority and KG Jebsen Stiftelsen. We further thank the Center for Diabetes Research, the University of Bergen for providing genotype data and performing quality control and imputation of the data funded by the ERC AdG project SELECTionPREDISPOSED, Stiftelsen Kristian Gerhard Jebsen, Trond Mohn Foundation, the Research Council of Norway, the Novo Nordisk Foundation, the University of Bergen, and the Western Norway health Authorities (Helse Vest). The authors thank FinnGen investigators for sharing their summary-level data.

## Notes

### Author Declarations

Ethical approval for UKB was obtained from the North West Multi-centre Research Ethics Committee, and our study was performed under UKB application number 23938. Ethical approval for the study was obtained from the ALSPAC Ethics and Law Committee and the Local Research Ethics Committees. Ethical approval for BiB was obtained from the Bradford Research Ethics Committee. The establishment of MoBa and initial data collection was based on a license from the Norwegian Data Protection Agency and approval from The Regional Committees for Medical and Health Research Ethics.

