## Supplement for "Evaluating causal associations of chronotype with pregnancy and perinatal outcomes and its interactions with insomnia and sleep duration: a Mendelian randomization study"

**eAppendix 1. Descriptions of each cohort**

***UK Biobank (UKB)***

All people in the UK National Health Service registry aged between 40-69 years and living within a 25 mile radius from one of 22 study centres were invited to participate between 2006-2010.^1^ In total 503 325 adults (5.5% of the ~9.2 million invited) were recruited into UKB.^1^ Participants who had a valid email address (N=339 229) were invited to fill in a detailed online questionnaire assessing their mental health after January 2015, and 158 835 participants fully completed it by October 2017.^2^ Ethical approval for UKB was obtained from the North West Multi-centre Research Ethics Committee, and our study was performed under UKB application number 23938.

Age (years) was derived based on date of birth and date of attending an initial assessment centre. Women were asked to report their age when they had their only one child, or the first child if they had given birth to more than one child. Offspring sex was not available at the initial assessment. Chronotype, insomnia and sleep duration were self-reported at UKB initial assessment centre. Chronotype was measured via a question – “Do you consider yourself to be?” with options “definitely a ‘morning’ person”, “more a ‘morning’ than ‘evening’ person”, “more an ‘evening’ than a ‘morning’ person”, “definitely an ‘evening’ person”, “do not know” and “prefer not to answer”. We derived a binary variable of chronotype by coding these options as 1, 1, 0, 0, missing and missing, respectively, consistent with the most updated GWAS.^3^ Insomnia was measured via a question “Do you have trouble falling asleep at night or do you wake up in the middle of the night?” with responses “never/rarely”, “sometimes”, “usually” and “prefer not to answer”. Insomnia was coded in the same way as the GWAS that provided genetic IVs (i.e. “usually” (cases) versus “sometimes” + “never/rarely” (controls)).^4^ Sleep duration was measured via a question “About how many hours sleep do you get in every 24 hours? (please include naps).” The answer could only contain integer values. Following a previous MR study,^5^ participants whose sleep duration ranged from 2 to 12 hours (206,500 women, 99.2%) were eligible to be included into our analyses.

Genotyping, pre-imputation quality control, and imputation procedures were described in detail elsewhere,^6^ and we briefly summarized here. UKB men and women were genotyped on two arrays. The first ~50 000 samples were genotyped on the UK BiLEVE array and the remaining ~450 000 samples were genotyped on the UK Biobank Axiom array. Genotype data were imputed against two references panels: Haplotype Reference Consortium (HRC) panel and UK10K + 1000 Genomes panel. We used the imputed data released by UKB in March 2018, and applied in-house post-imputation quality controls (QC, i.e. genetic sex same as reported sex, XX or XY in sex chromosome and no outliers in heterozygosity and missing rates).^7^ Women of European descent with qualified genotype data were eligible for inclusion in our Mendelian randomization (MR) analyses (N=176 897, see eFigure 8A in Supplement).

***Avon Longitudinal Study of Parents and Children (ALSPAC)***

Pregnant women resident in Avon, UK with expected dates of delivery 1^st^ April 1991 to 31^st^ December 1992 were invited to take part in the study.^8,9^ The initial number of pregnancies enrolled is 14 541 (for these at least one questionnaire has been returned or a ‘Children in Focus’ clinic data had been attended by 19/07/99).^8,9^ Of these initial pregnancies, there was a total of 14 676 foetuses, resulting in 14 062 live births and 13 988 children who were alive at 1 year of age.^8,9^ Our study relied on 13 867 pregnancies from a total of 13 761 women, and most pregnancies were recruited at the first antenatal clinic visit in the first trimester of pregnancy.^8^ Questionnaires were sent at a regular interval during pregnancy, and biological samples were taken from parents and children including blood samples from which DNA was extracted. Please note that the study website (<http://www.bristol.ac.uk/alspac/researchers/our-data/>) contains details of all the data that is available through a fully searchable data dictionary and variable search tool. Ethical approval for the study was obtained from the ALSPAC Ethics and Law Committee and the Local Research Ethics Committees. Consent for biological samples has been collected in accordance with the Human Tissue Act (2004). Informed consent for the use of data collected via questionnaires and clinics was obtained from participants following the recommendation of the ALSPAC Ethics and Law Committee at the time.

Maternal age at delivery (years) was derived from date of delivery and date of birth. Offspring sex for the index pregnancy was provided by ALSPAC researchers, according to multiple sources (including obstetric data, birth notifications or clinical records). Insomnia in pregnancy was self-reported, at 18 weeks of gestation, using the question “Can you get off to sleep alright?” with options “Very often,” “Often,” “Not very often,” and “Never.” We compared “Not very often” + “Never” [i.e. insomnia cases] versus “Very often” + “Often” [i.e. no insomnia].

Genotyping, pre-imputation quality control, and imputation procedures were described in detail elsewhere,^10^ and we briefly summarized here. ALSPAC mothers were genotyped using Illumina human660K quad SNP chip, and ALSPAC children were genotyped using Illumina HumanHap550 quad genome-wide SNP genotyping platform. Genotype data for both ALSPAC mothers and children were imputed against HRC v1.1 reference panel, after a similar QC procedure (minor allele frequency (MAF) ≥1%, a call rate ≥95%, in Hardy-Weinberg equilibrium (HWE), correct sex assignment, no evidence of cryptic relatedness, and of European decent). Women of European descent with qualified genotype data and live-born singleton offspring were eligible for inclusion in our MR analyses (N=6826, see eFigure 8B in Supplement).

***Born in Bradford (BiB)***

BiB is a birth cohort that recruited 13 776 pregnancies to 12 453 women resident in the Bradford metropolitan district (a city in the North of England), with expected dates of delivery between 2007-2011.^11^ Most pregnancies were recruited during an oral glucose tolerance test undertaken between 24-28 weeks of gestation, and to which all pregnant women in Bradford were invited.^11^ BiB reflects the multicultural mix profile of Bradford, with approximately 50% of the obstetric population being of South Asian descent.^11^ The study website (<https://borninbradford.nhs.uk/>) provides details of all available data. Ethical approval for BiB was obtained from the Bradford Research Ethics Committee. Information on maternal age (years) was collected via a questionnaire at baseline. Offspring sex was extracted from maternity electronic record by BiB research team.

Genotyping, pre-imputation quality control, and imputation procedures were described elsewhere,^12^ and we briefly summarized here. Both BiB mothers and BiB children were genotyped using Illumina HumanCoreExome chip. Genotype data for both of them were imputed against UK10K + 1000 Genomes reference panel, after a similar QC procedure (a call rate ≥99.5%, correct sex assignment, no evidence of cryptic relatedness, correct ethnicity assignment). Two subsets of individuals were declared based on a combination of principal component analysis and self-report ethnicity. To combine the MR estimates with those in other cohorts, only women of European descent with qualified genotype data and live-born singleton offspring were eligible for inclusion in our analyses (N=2940, see eFigure 8C in Supplement).

***Norwegian Mother and Child Cohort Study (MoBa)***

MoBa is a population-based pregnancy cohort study conducted by the Norwegian Institute of Public Health. Participants were recruited from all over Norway from 1999-2008.^13^ The women consented to participation in 41% of the pregnancies. The cohort now includes 114,500 children, 95,200 mothers and 75,200 fathers.^13^ The current study is based on version 12 of the quality-assured data files released for research in “Prenatal environmental exposures and pregnancy outcomes-Mendelian randomization analysis”. The establishment of MoBa and initial data collection was based on a license from the Norwegian Data Protection Agency and approval from The Regional Committees for Medical and Health Research Ethics. The MoBa cohort is now based on regulations related to the Norwegian Health Registry Act. The current study was approved by The Regional Committees for Medical and Health Research Ethics. The Medical Birth Registry (MBRN) is a national health registry containing information about all births in Norway. MoBa has been linked to the Medical Birth Register of Norway (MBRN, established in 1967), using unique personal identification numbers.^13^

Maternal age at delivery and offspring sex were recorded in Medical Birth Registry of Norway. Sleep duration was assessed via a self-administered question—“How many hours a day do you usually sleep now when you are pregnant?” at 30 weeks of gestation. Women reported their sleep duration in five categories, which were “over 10 h”, “8–9 h”, “6–7 h”, “4–5 h” and “less than 4 h”. The questionnaire did not specify whether to include naps so it is unclear whether the women would have reported duration only for night sleep or across 24 h (as in UKB). Due to small numbers, we combined the last two categories into “≤5 h”.

Blood samples were obtained from both parents during pregnancy and from mothers and children (umbilical cord) at birth.^14^ Genotyping, pre-imputation quality control, and imputation procedures were described in detail elsewhere,^15,16^ and we briefly summarized here. There were five projects contributed to MoBa genetics 1.0.^17^ In HARVEST, participants were genotyped using either Illumina HumanCoreExome12v1.1 or Illumina HumanCoreExome24v1.0. In ROTTERDAM1 & 2, participants were genotyped using Illumina GSAMDv1.0. In NORMENT1, participants were genotyped by 4 sub-projects with different batches. In TED, participants were genotyped using Illumina InfiniumOmniExpress-24v1-2. Genotype data from all batches were imputed against HRC v1.1 and IMPUTE4 reference panels, after a similar QC procedure (MAF ≥0.5%, a call rate ≥95%, in HWE, correct sex assignment and no evidence of cryptic relatedness). Women of European descent with qualified genotype data and live-born singleton offspring were eligible for inclusion in our analyses (N=57 430, see eFigure 8D in Supplement).

***FinnGen***

FinnGen is the national wide network of Finnish biobanks, including Auria Biobank, Biobank Borealis of Northern Finland, Biobank of Eastern Finland, Central Finland Biobank, Finnish Red Cross Blood Service Biobank, Finnish Clinical Biobank Tampere, Helsinki Biobank, Terveystalo Biobank, and THL Biobank.^18^ Those biobanks were linked to national registries, including Drug purchase and Drug Reimbursement, Digital and Population Data Services Agency, Statistics Finland, Register of primary health care visits: AVOHILMO, Care Register for Health Care: HILMO, and Finnish cancer registry. Clinical endpoints were defined based on ICD-10, and the equivalent in ICD-8 and ICD-9. The Coordinating Ethics Committee of the Helsinki and Uusimaa Hospital District has approved the FinnGen consortium (Nr HUS/990/2017), and the ethical approval of each individual study has been described in detail elsewhere.^19^ FinnGen participants were genotyped with Illumina and Affymetrix chip arrays. Genotype data was imputed against SISu v3 reference panel (<http://sisuproject.fi>), after a QC procedure (minor allele count ≥3, a call rate ≥98%, in HWE no outliers in heterozygosity, correct sex assignment, and of Finnish ancestry). Full details of FinnGen genotype data were summarized elsewhere.^18^

**eAppendix 2. Two-sample Mendelian randomization (MR) stratified on genetically predicted chronotype**

We calculated SNP-exposure associations by fitting logistic regression for insomnia^20^ and linear regression for sleep duration,^21^ adjusting for genotyping batch, top 10 principal components (PCs) and women’s age in UKB. We calculated SNP-outcome associations by fitting logistic regression (linear regression for offspring birthweight), adjusting for top 10 PCs and women’s age (and genotyping batches in UKB and MoBa where multiple batches were used).

Among women below and above the median, we obtained two-sample MR IVW estimates of insomnia and sleep duration on each outcome (Figure 1). In UKB, we followed previous studies to conduct a split cross-over two-sample MR.^5,20,21^ This meant that we used SNP-exposure associations from dataset A and SNP-outcomes associations from dataset B (A on B) and vice-versa (B on A). SNP-exposure associations, which were opposite to the original GWAS^4^ given fewer participants in the split samples, were excluded from two-sample MR analyses. We then meta-analysed the MR estimates from the two together for each exposure-outcome pair using fixed-effects (with inverse variance weights). For two-sample MR using ALSPAC, BiB and MoBa, we used SNP-exposure associations from UKB women and the pooled SNP-outcome associations combining the three birth cohorts. Finally, for each exposure-outcome pair, we pooled MR estimates from all four cohorts using fixed-effects (with inverse variance weights). eFigure 1 in Supplement illustrates our process.

**eFigure 1. Histogram of the GRS for evening preference in UK Biobank, ALSPAC, BiB and MoBa**

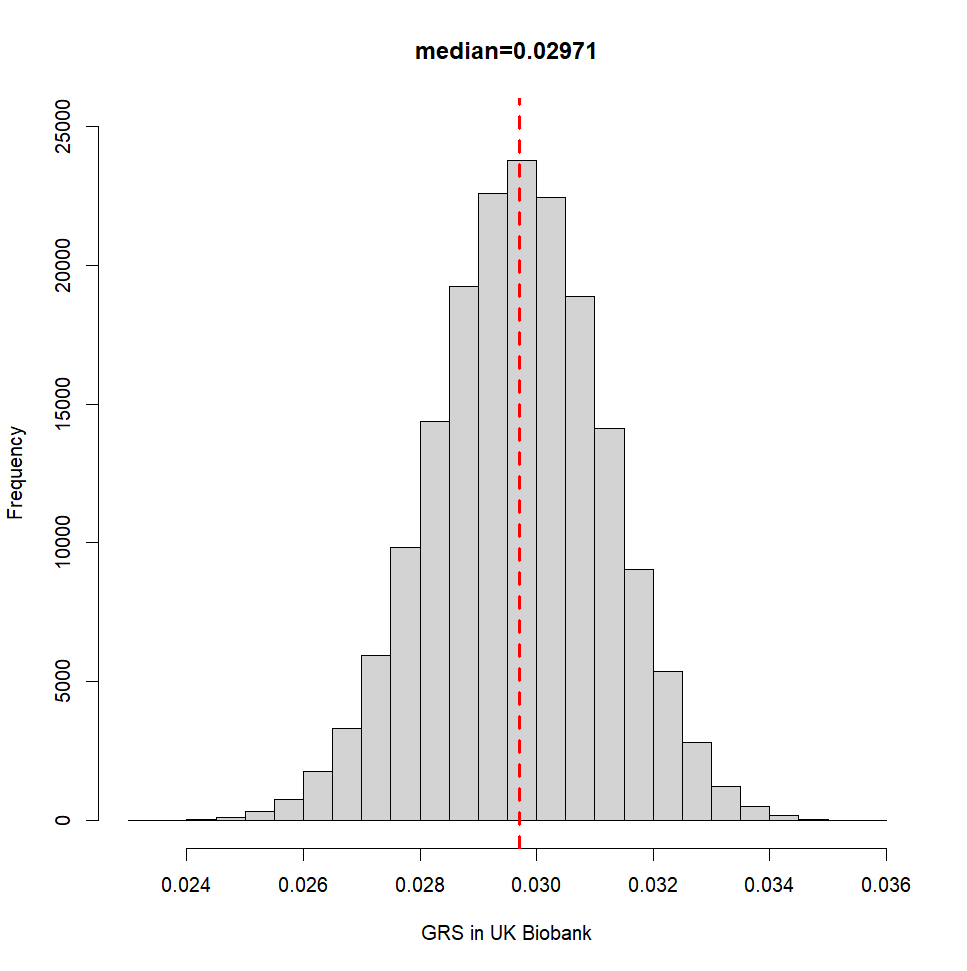

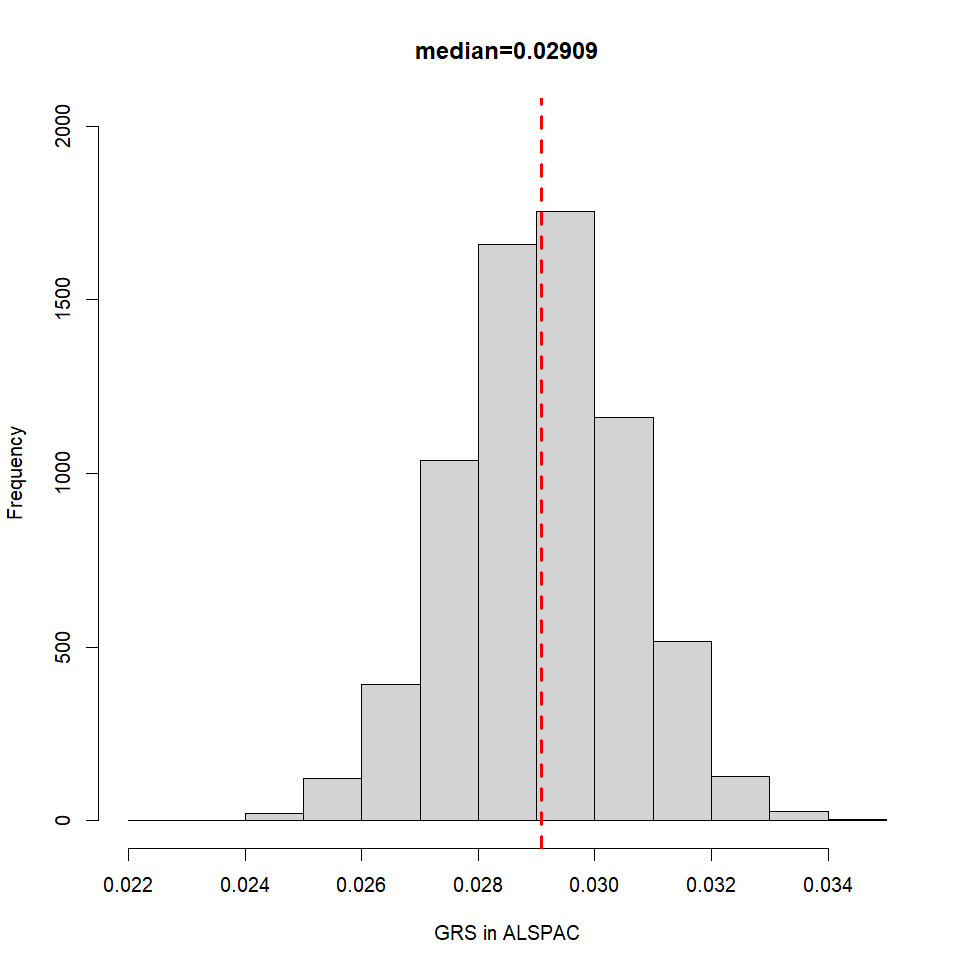

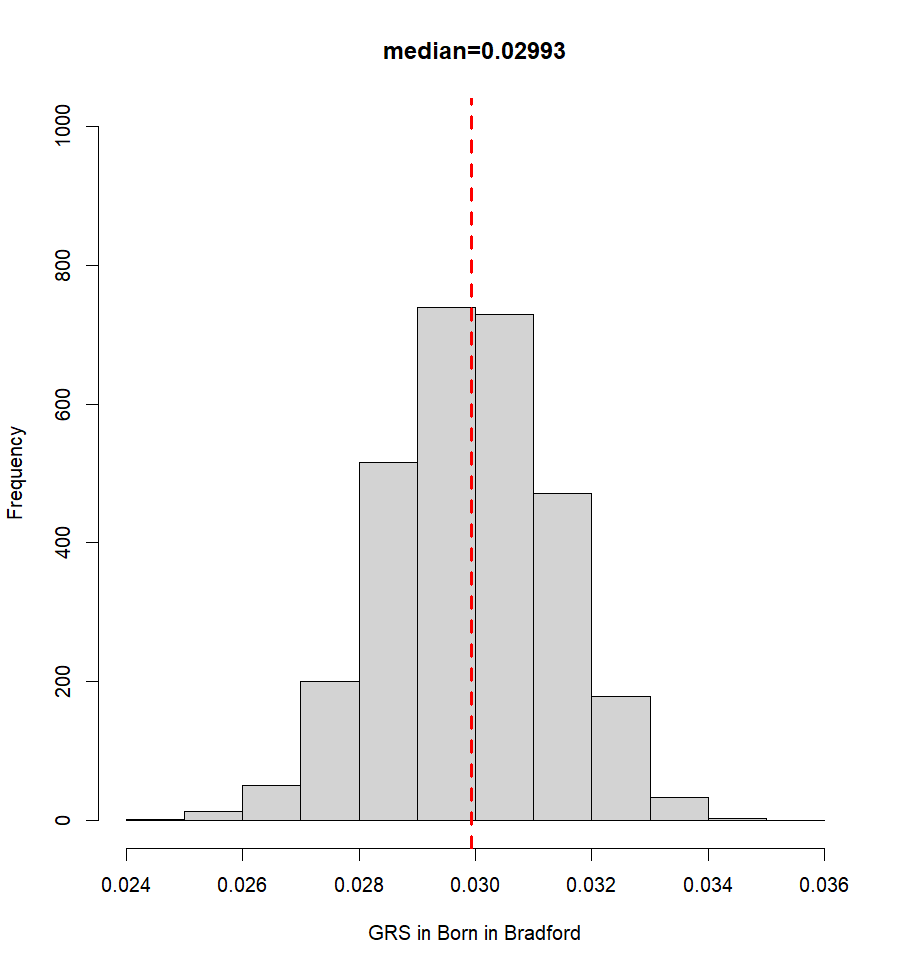

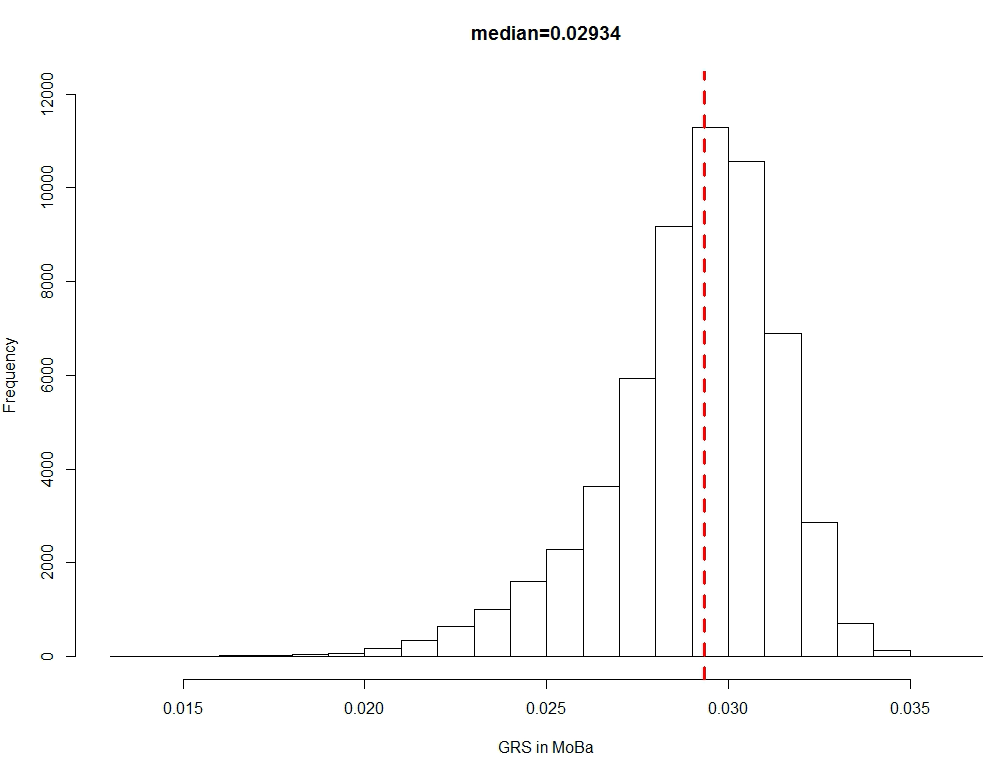

Abbreviations: ALSPAC, Avon Longitudinal Study of Parents and Children; GRS, genetic risk score; MoBa, Norwegian Mother, Father and Child Cohort Study.

**eFigure 2. Summary of methods and data contributing to two-sample MR stratified on genetically predicted chronotype**

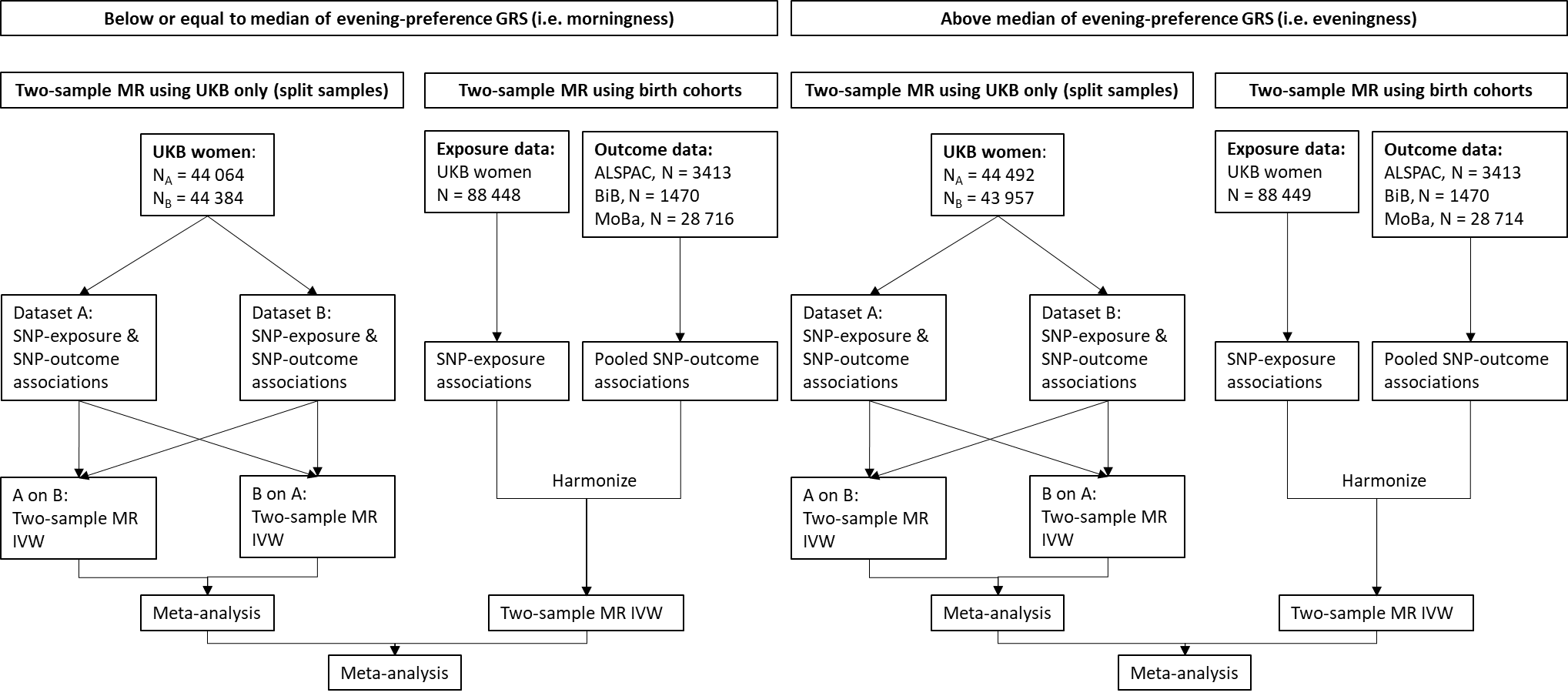

Abbreviations: ALSPAC, Avon Longitudinal Study of Parents and Children; BiB, Born in Bradford; GRS, genetic risk score; IVW, inverse variance weighted; MoBa, Norwegian Mother, Father and Child Cohort Study; MR, Mendelian randomization; SNP, single nucleotide polymorphism; UKB, UK Biobank.

**eFigure 3. Associations of 105 genome-wide significant single nucleotide polymorphisms with eveningness in 23andMe versus UK Biobank (UKB)**

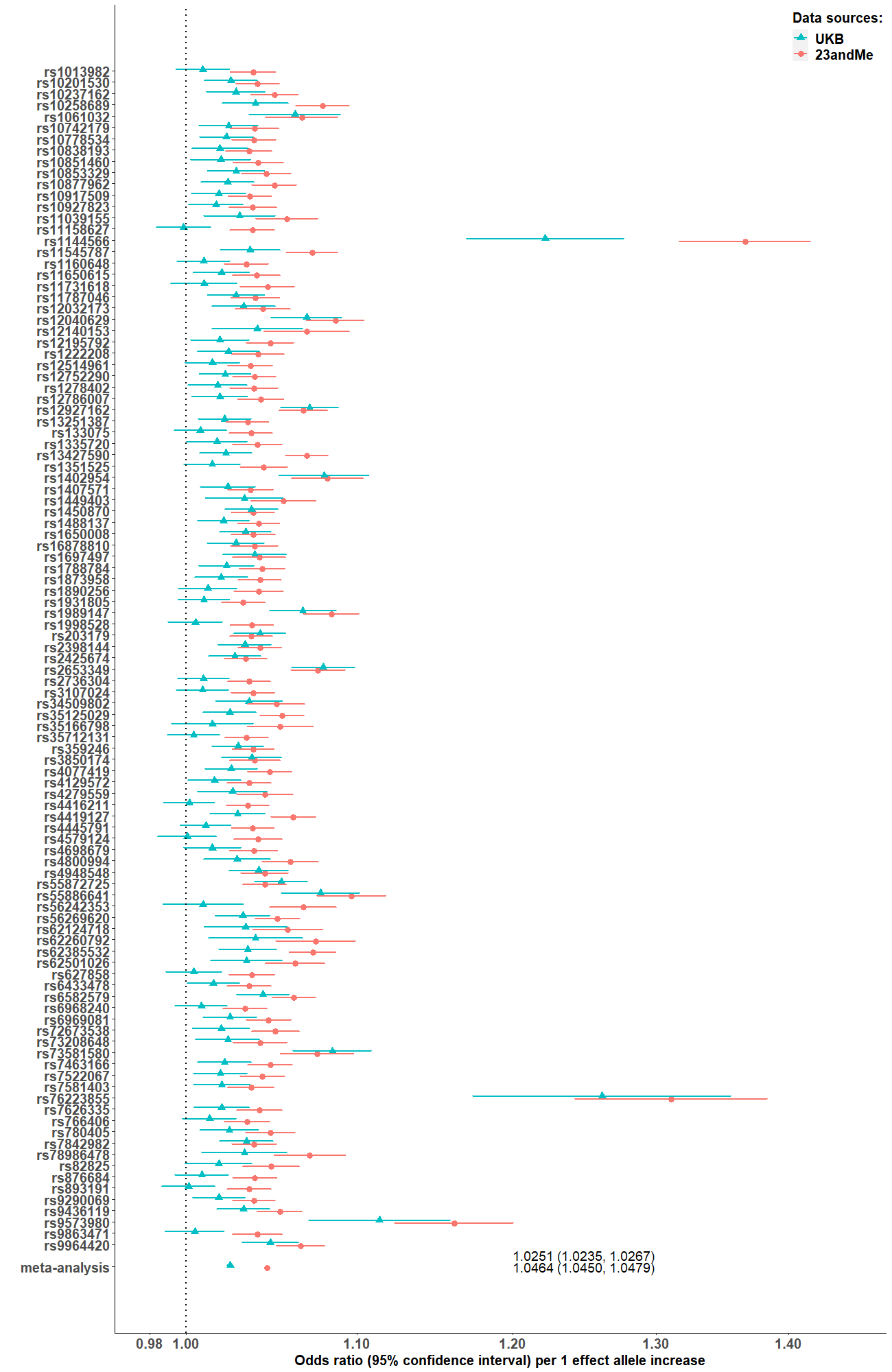

**eFigure 4. Leave-one study-out analyses for two-sample Mendelian randomization estimates of causal effects of chronotype on pregnancy and perinatal outcomes using inverse variance weighted**

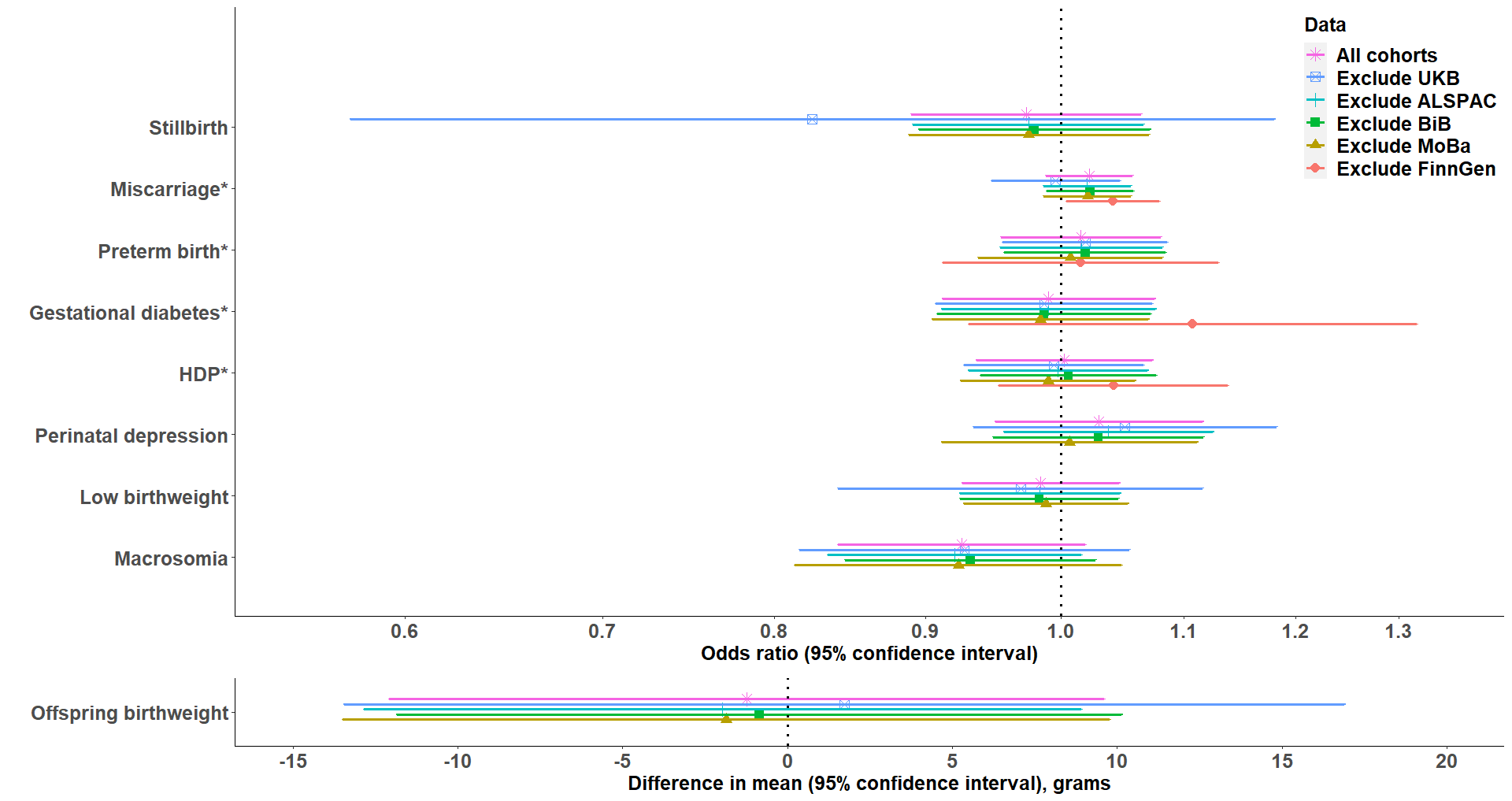

*FinnGen data were available for these outcomes.

Abbreviations: ALSPAC, Avon Longitudinal Study of Parents and Children; BiB, Born in Bradford; HDP, hypertensive disorders of pregnancy; MoBa, Norwegian Mother, Father and Child Cohort Study; UKB, UK Biobank.

**eFigure 5. Forest plots of two-sample MR estimates for causal effects of chronotype on pregnancy and perinatal outcomes using each single nucleotide polymorphism as an instrumental variable**

A. Stillbirth

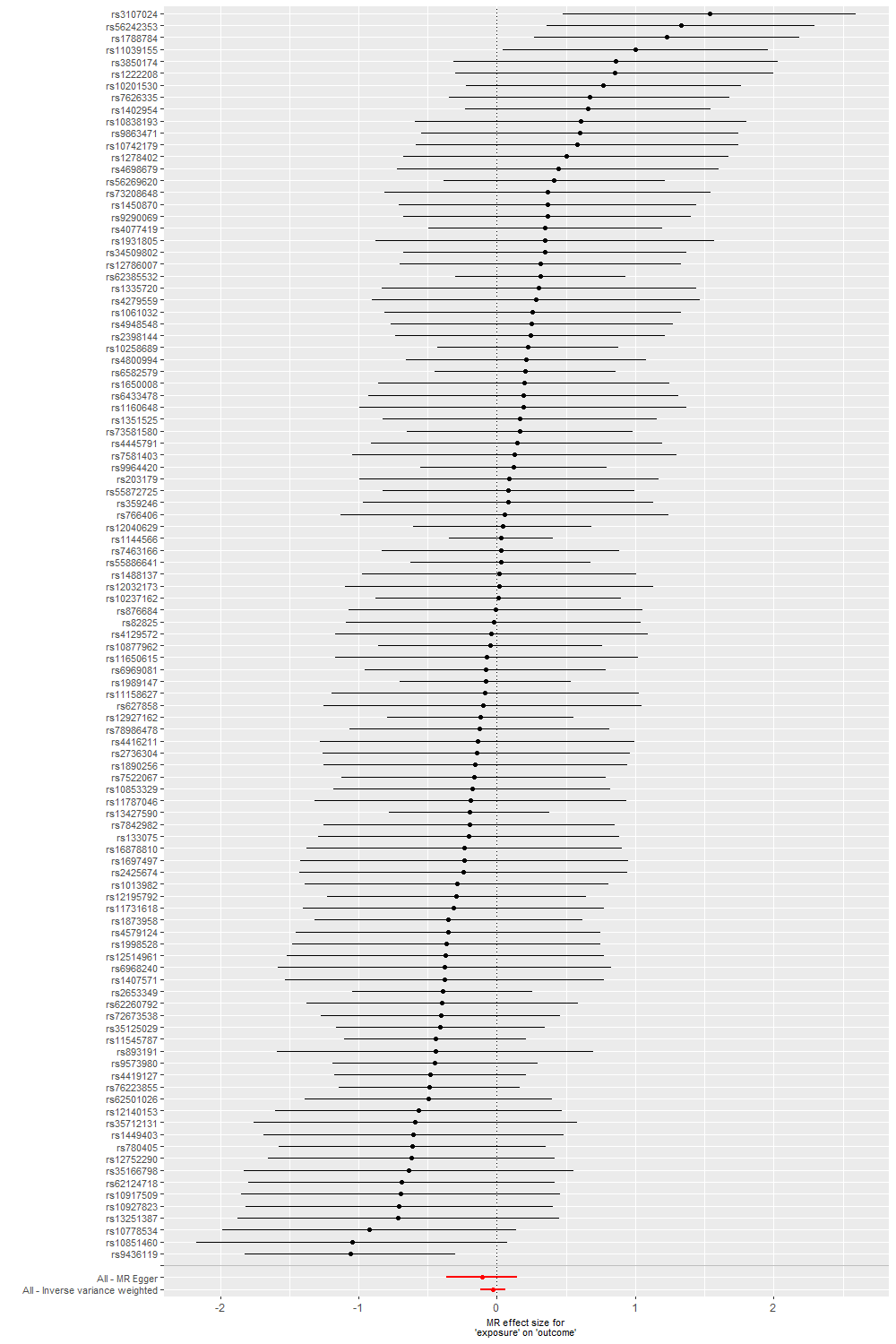

B. Miscarriage

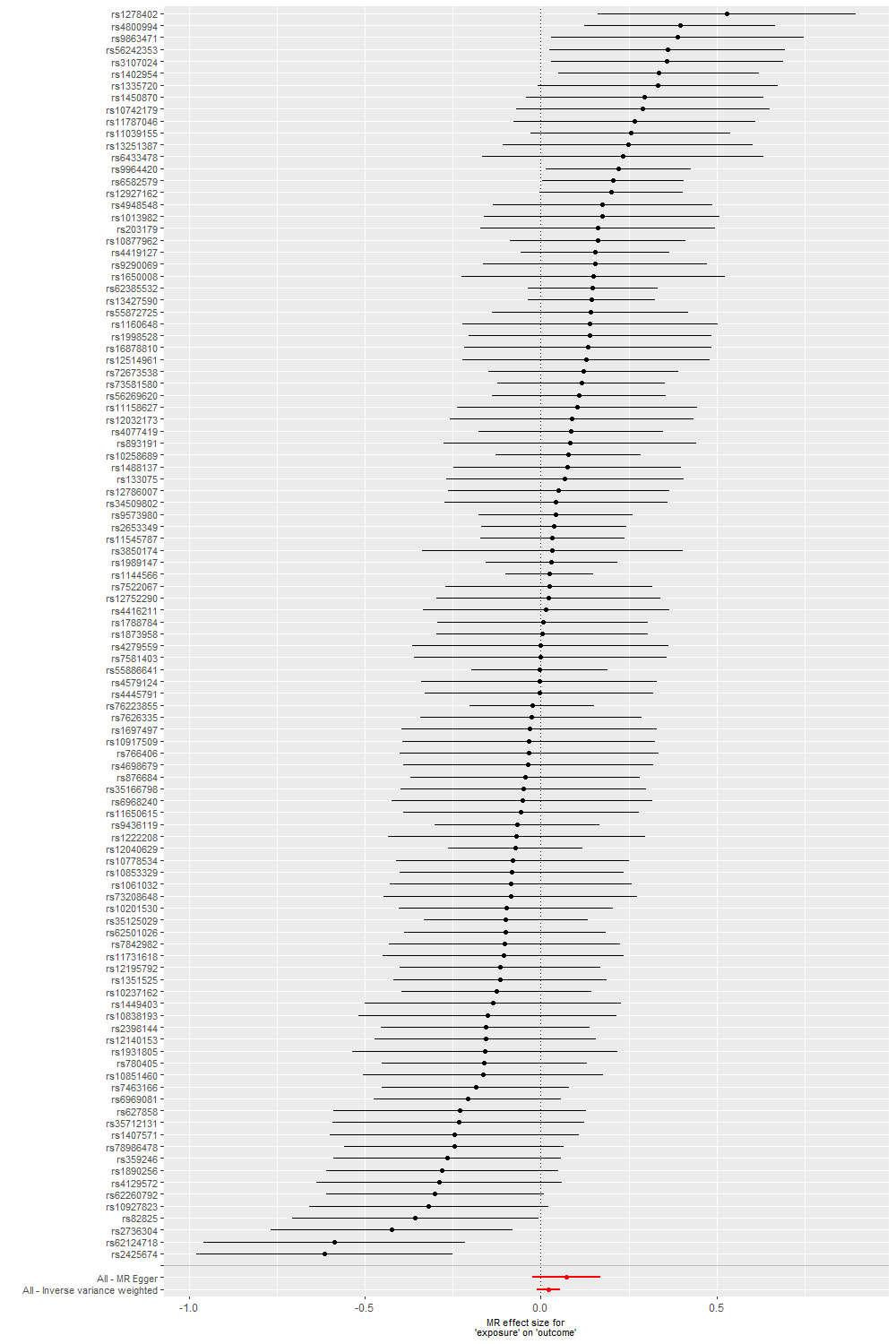

C. Preterm birth

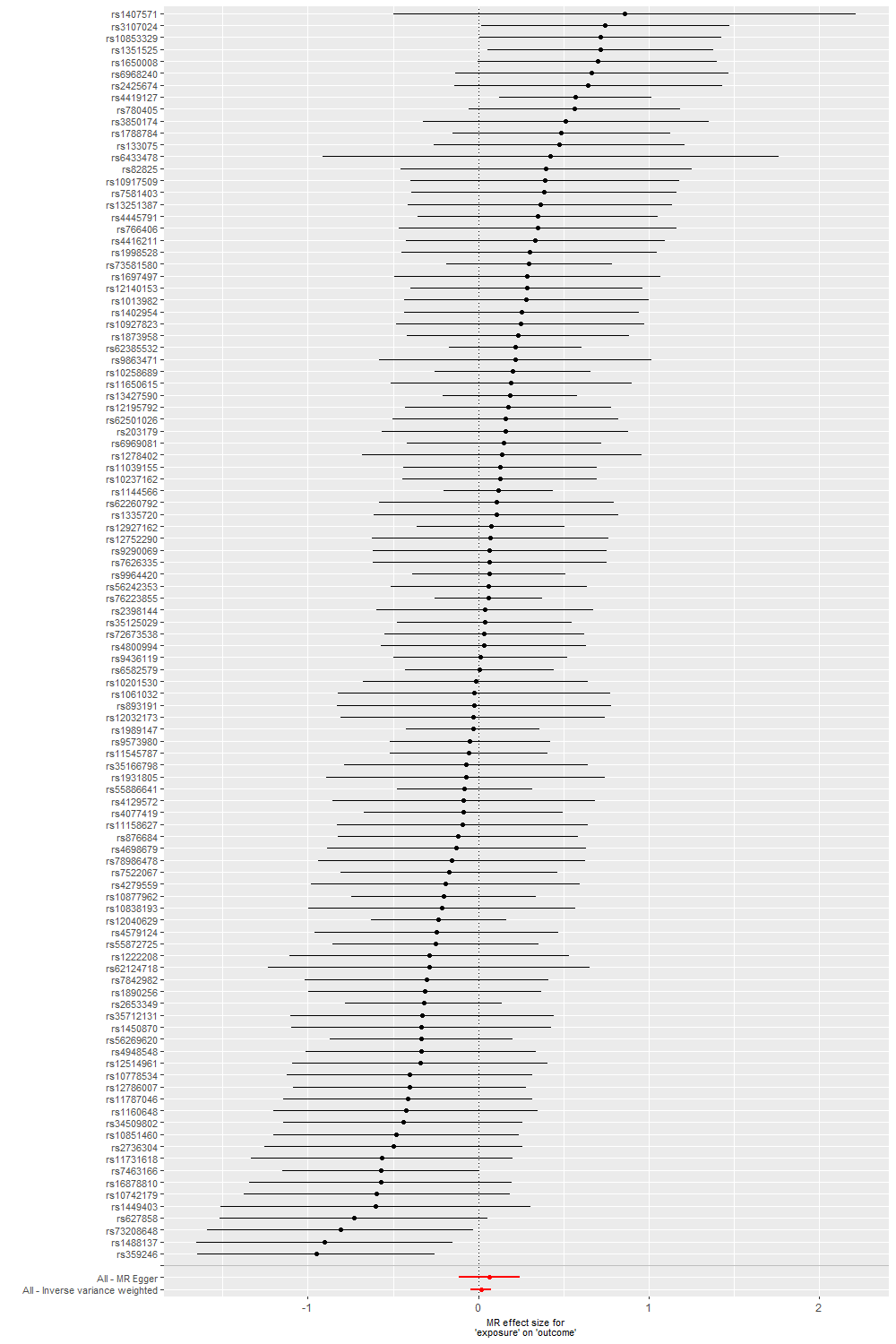

D. Gestational diabetes

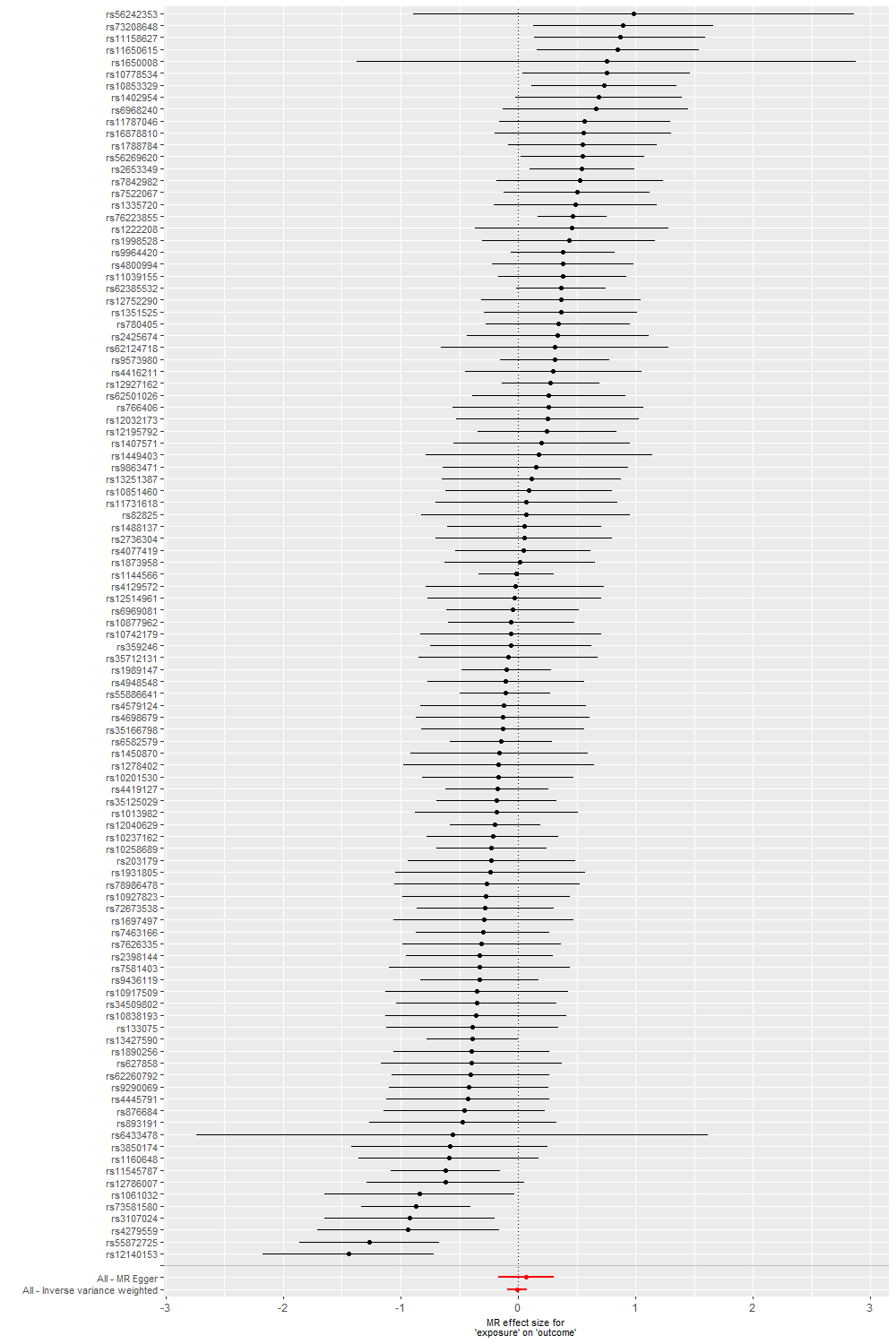

E. Hypertensive disorders of pregnancy

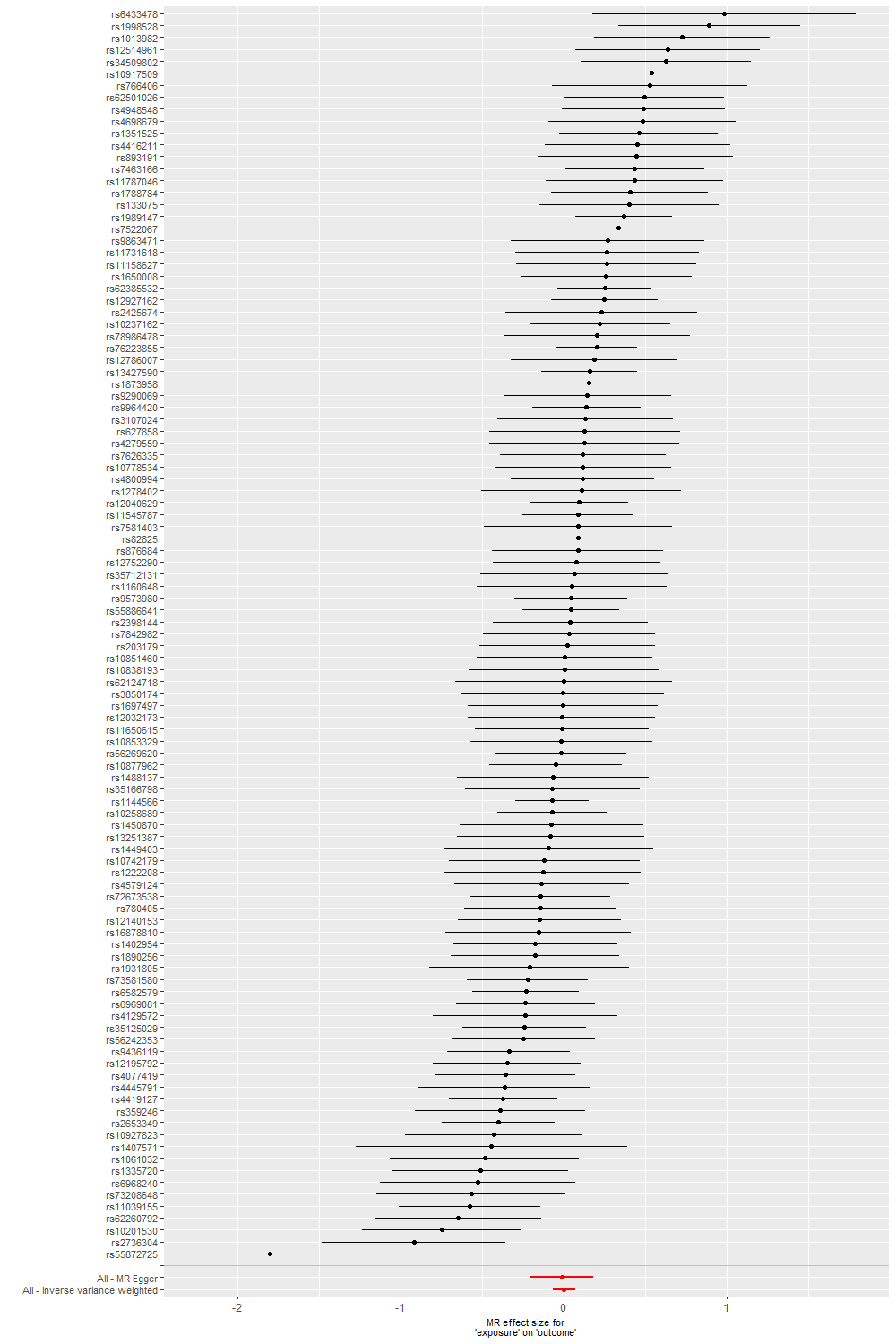

F. Perinatal depression

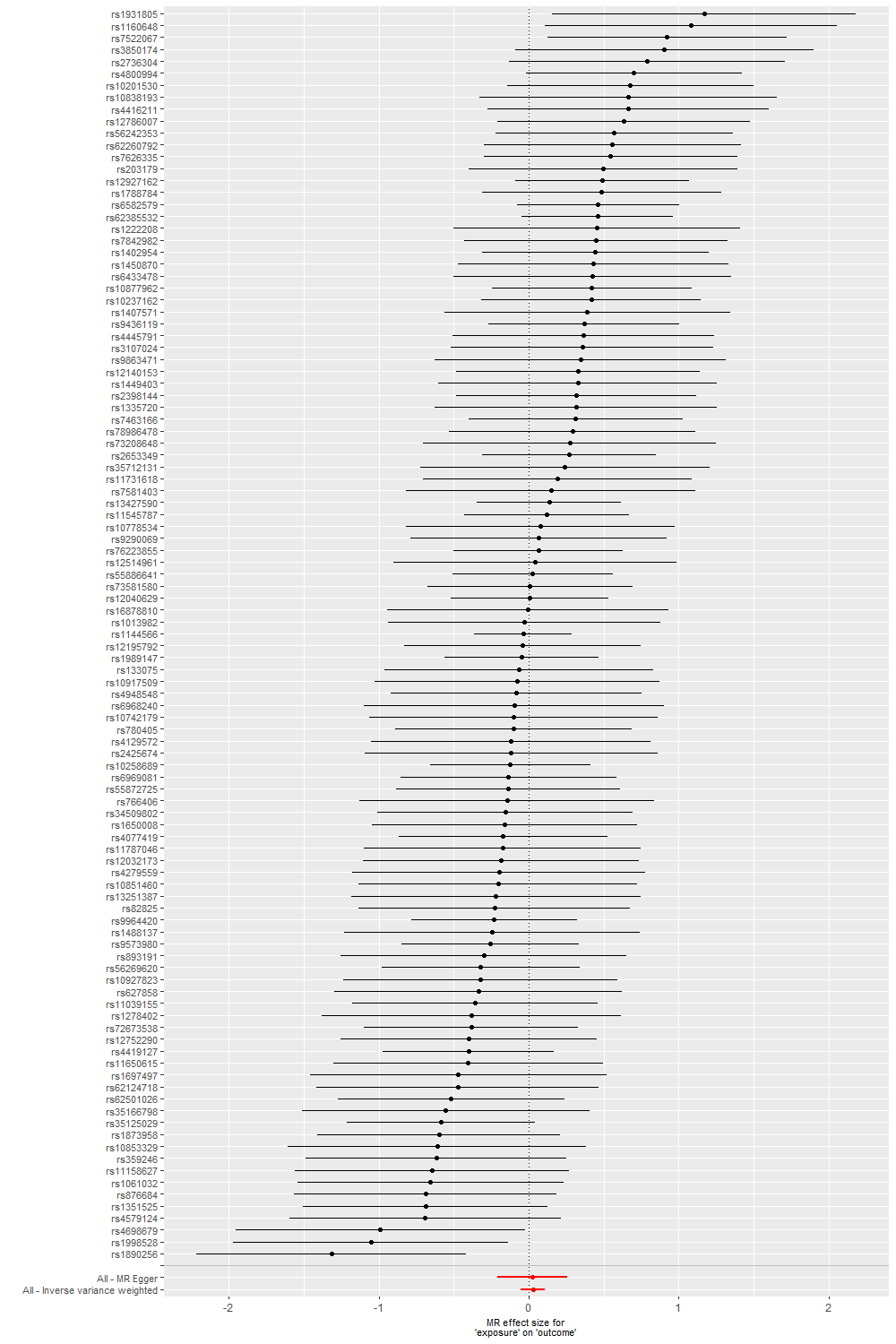

G. Low offspring birthweight

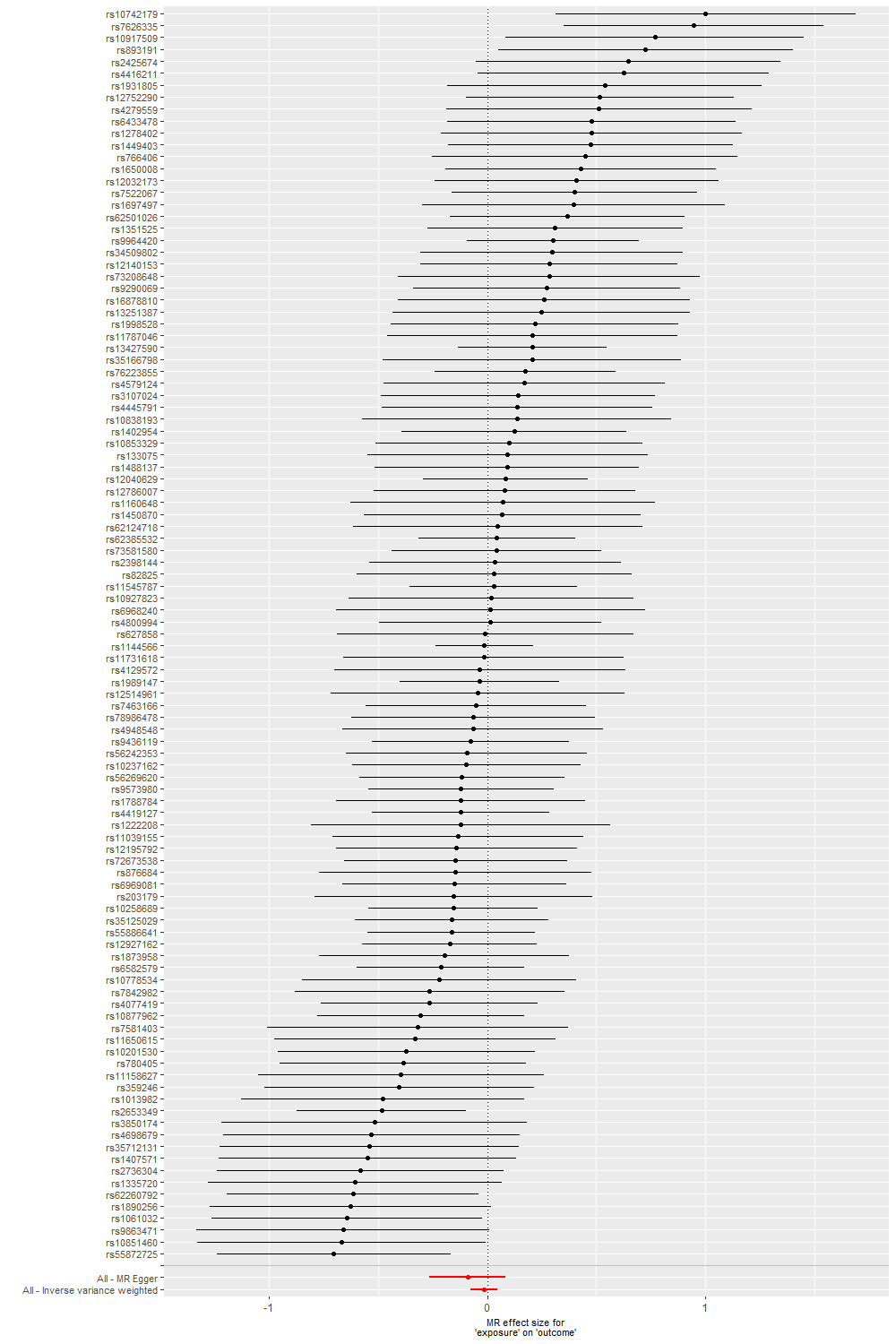

H. Macrosomia

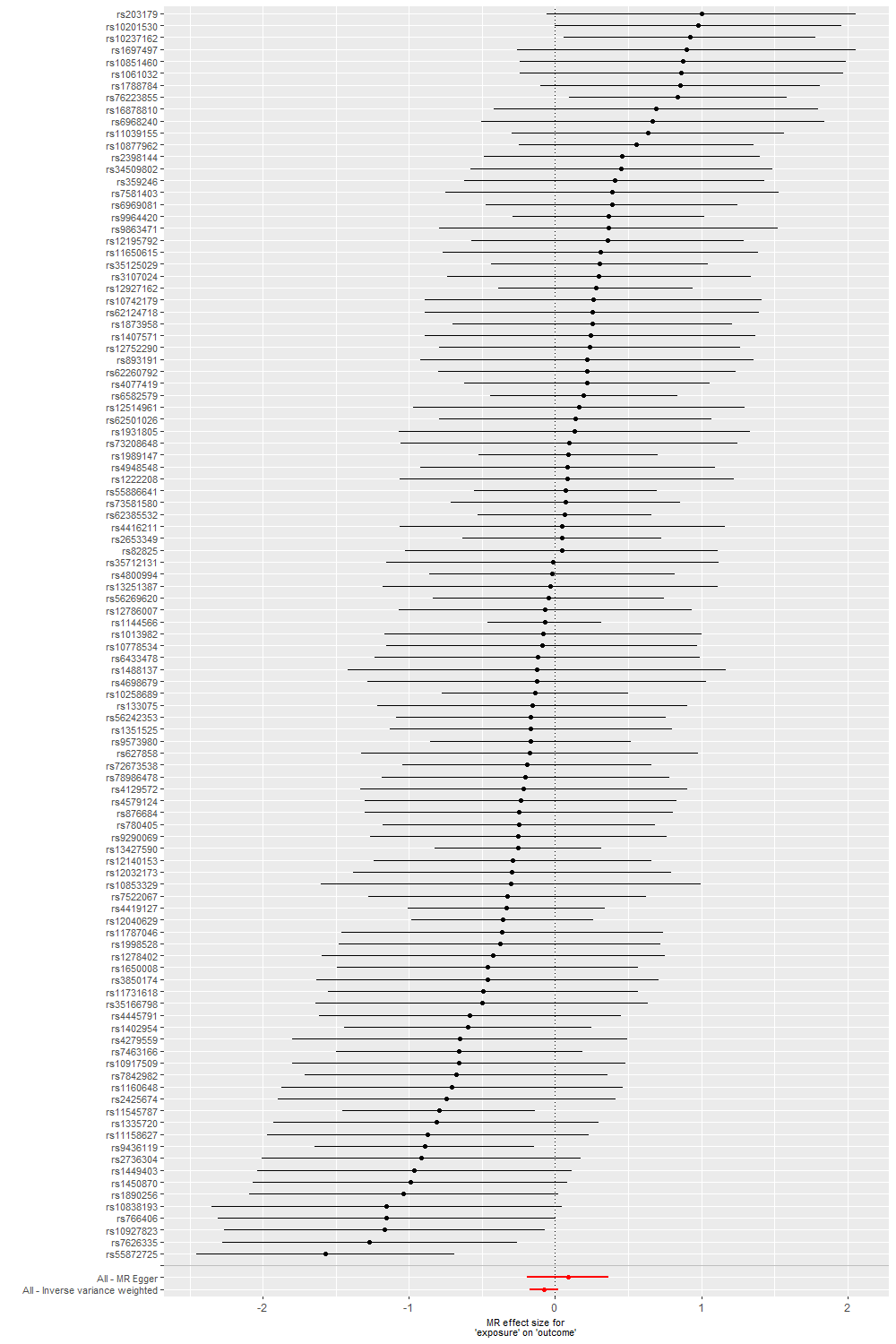

I. Offspring birthweight (grams)

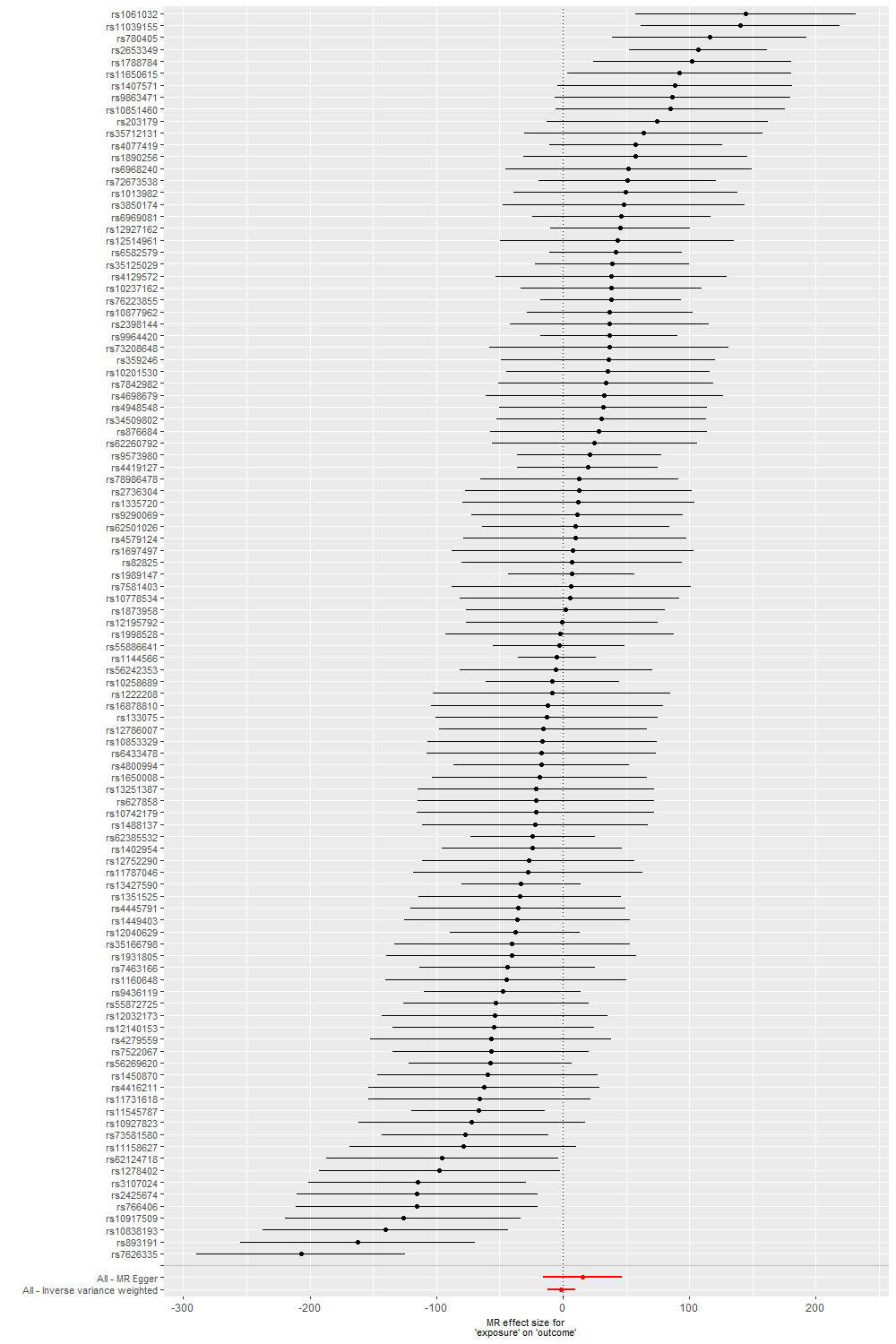

Abbreviation: MR, Mendelian randomization.

**eFigure 6. Leave-one SNP-out analyses for chronotype on pregnancy and perinatal outcomes in two-sample Mendelian randomization meta-analysing UK Biobank, ALSPAC, BiB, MoBa, and FinnGen**

A. Stillbirth

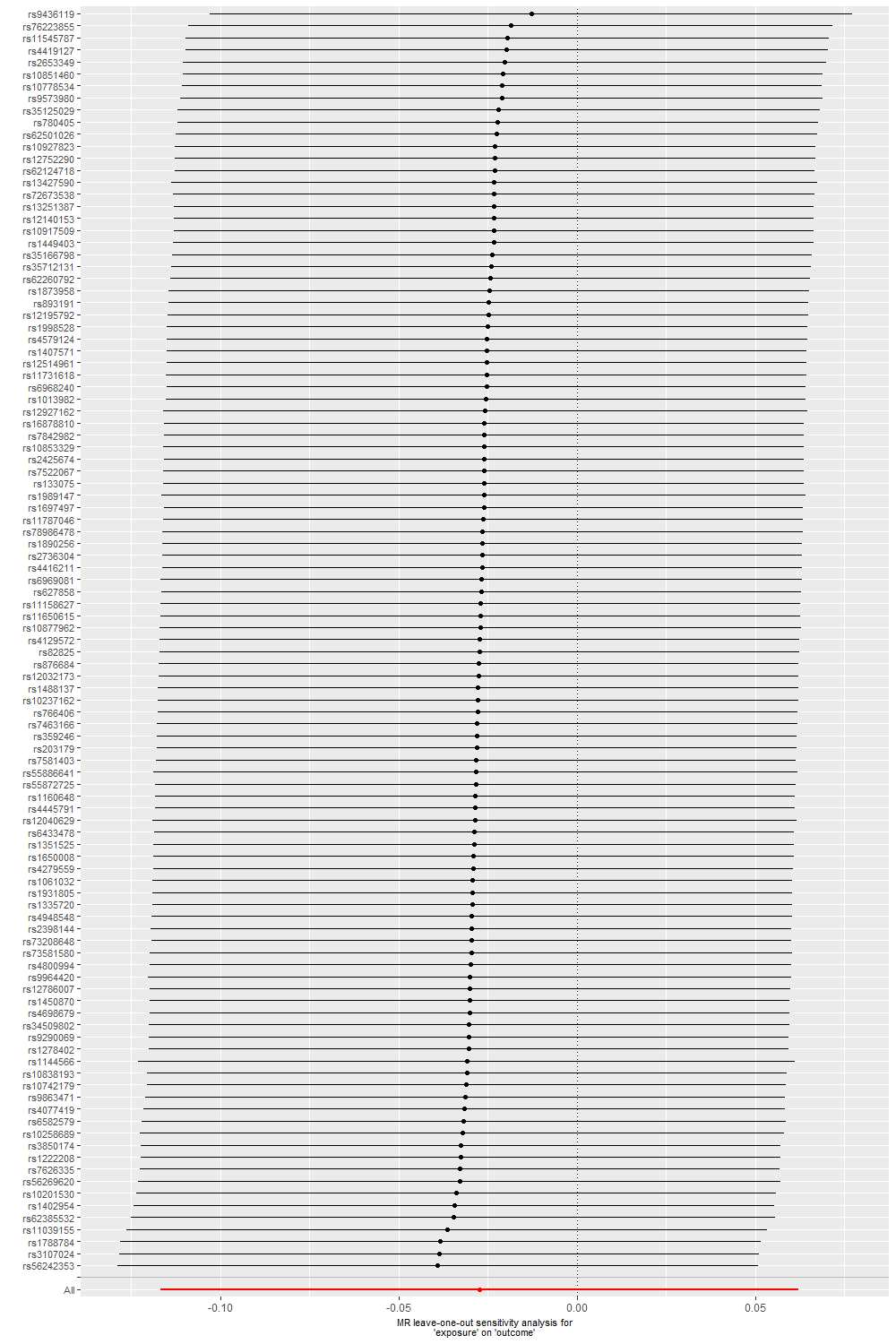

B. Miscarriage

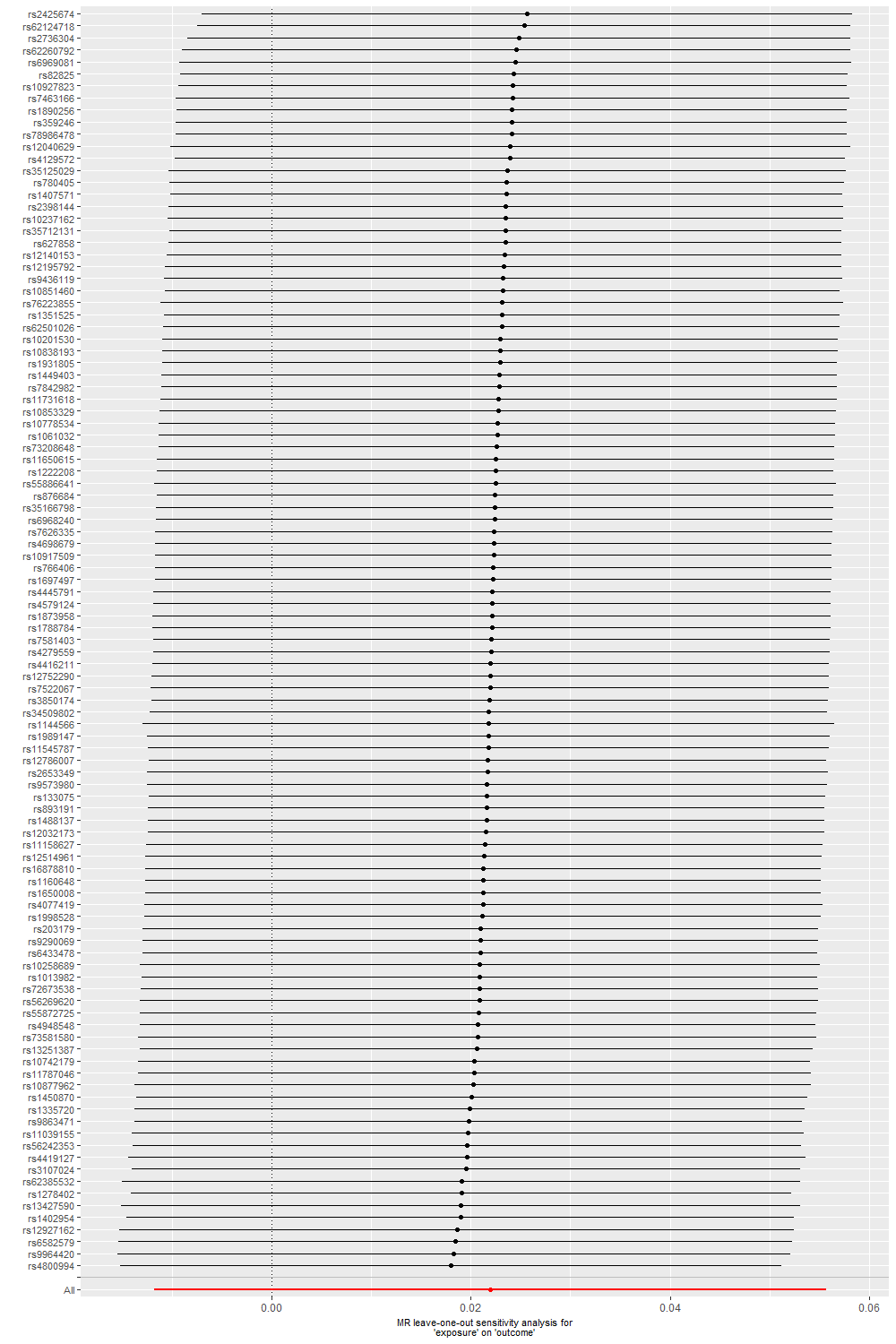

C. Preterm birth

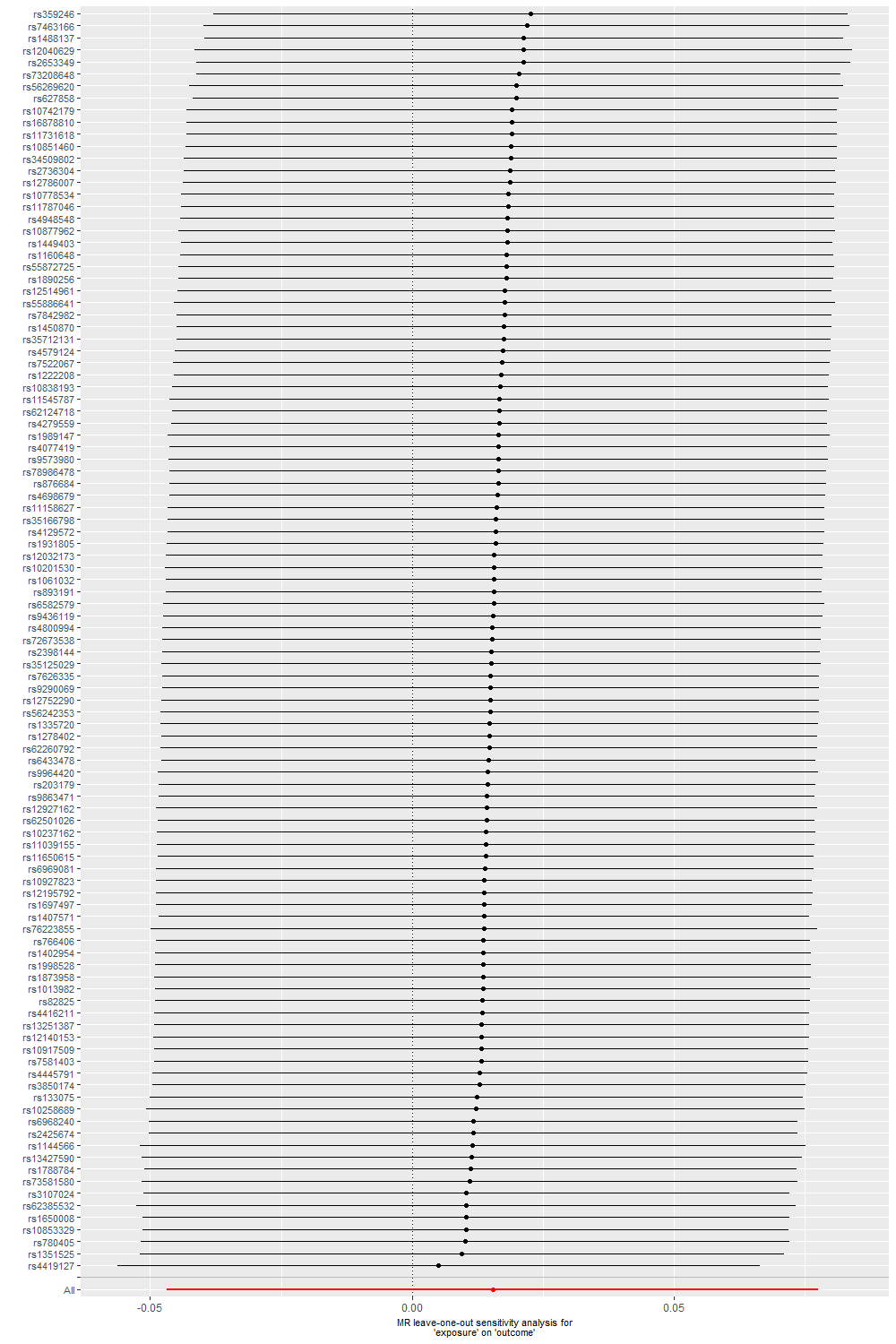

D. Gestational diabetes

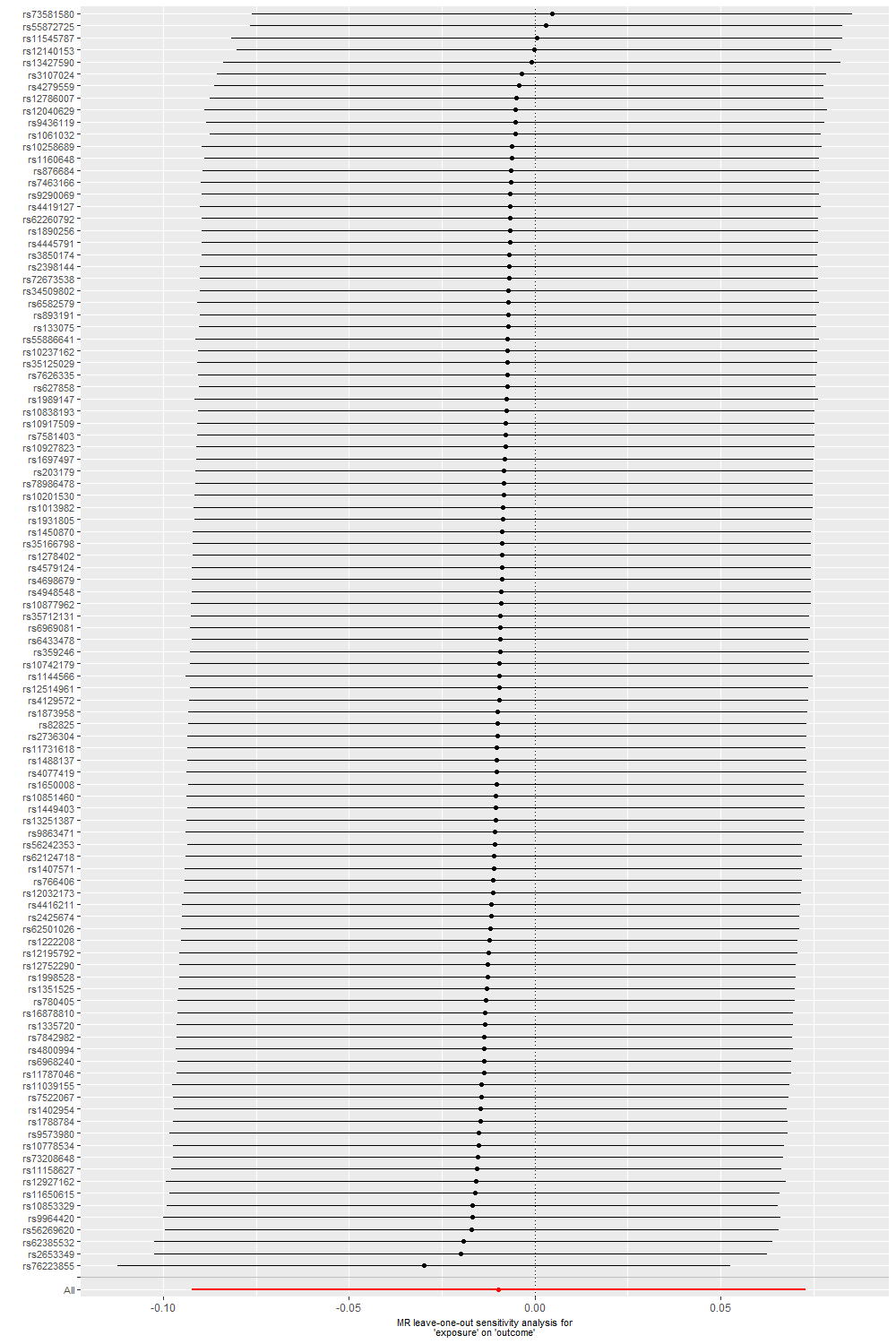

E. Hypertensive disorders of pregnancy

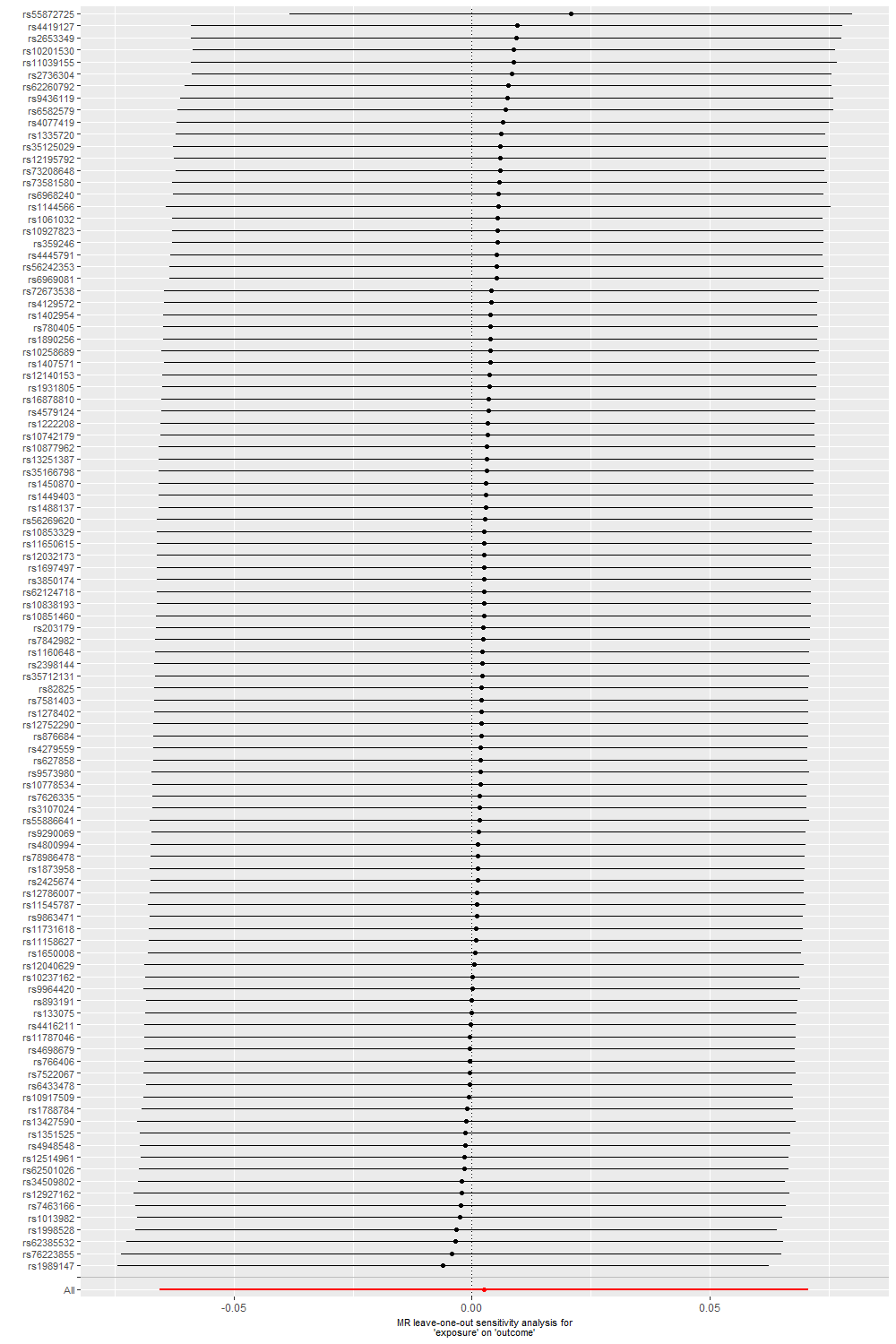

F. Perinatal depression

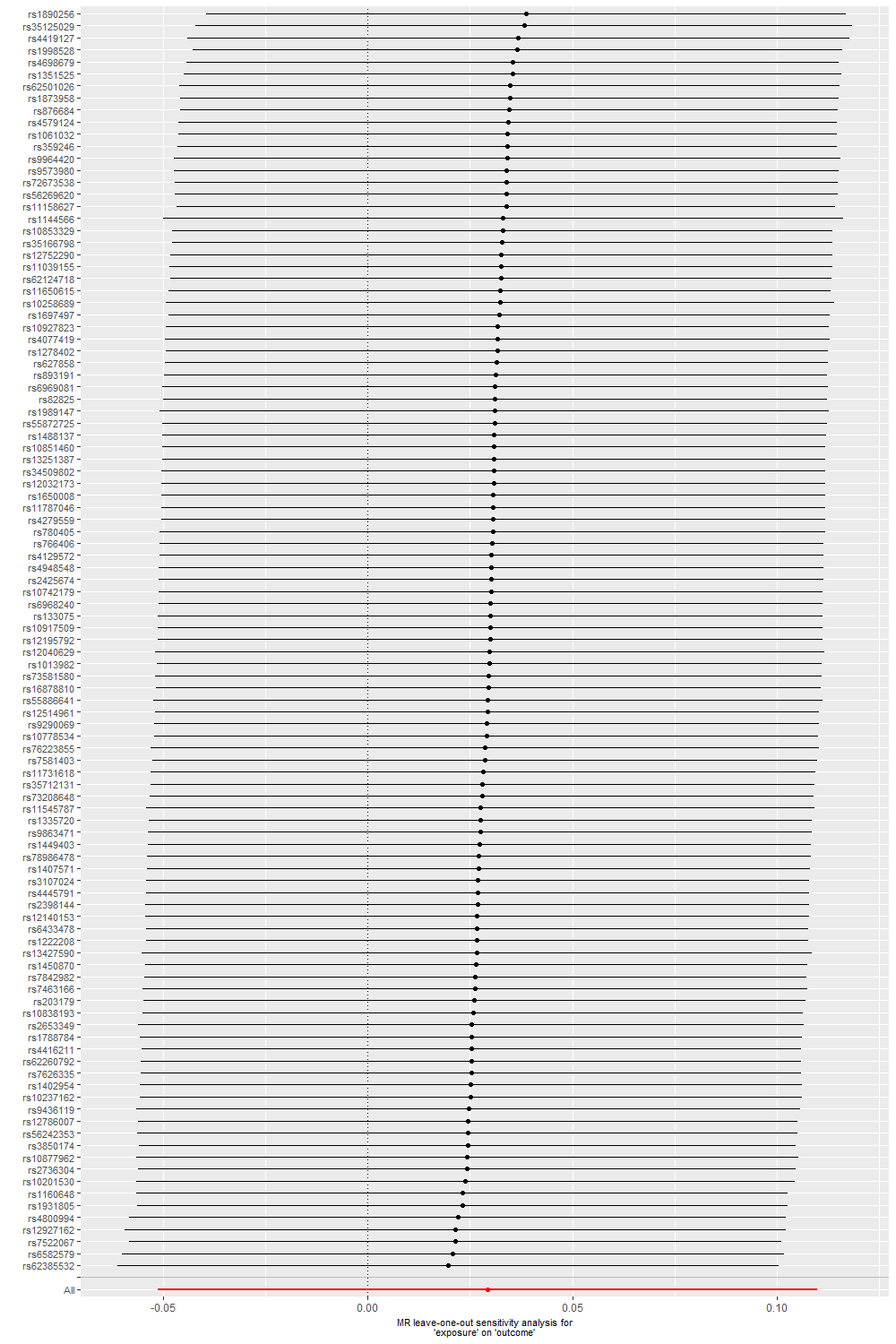

G. Low offspring birthweight

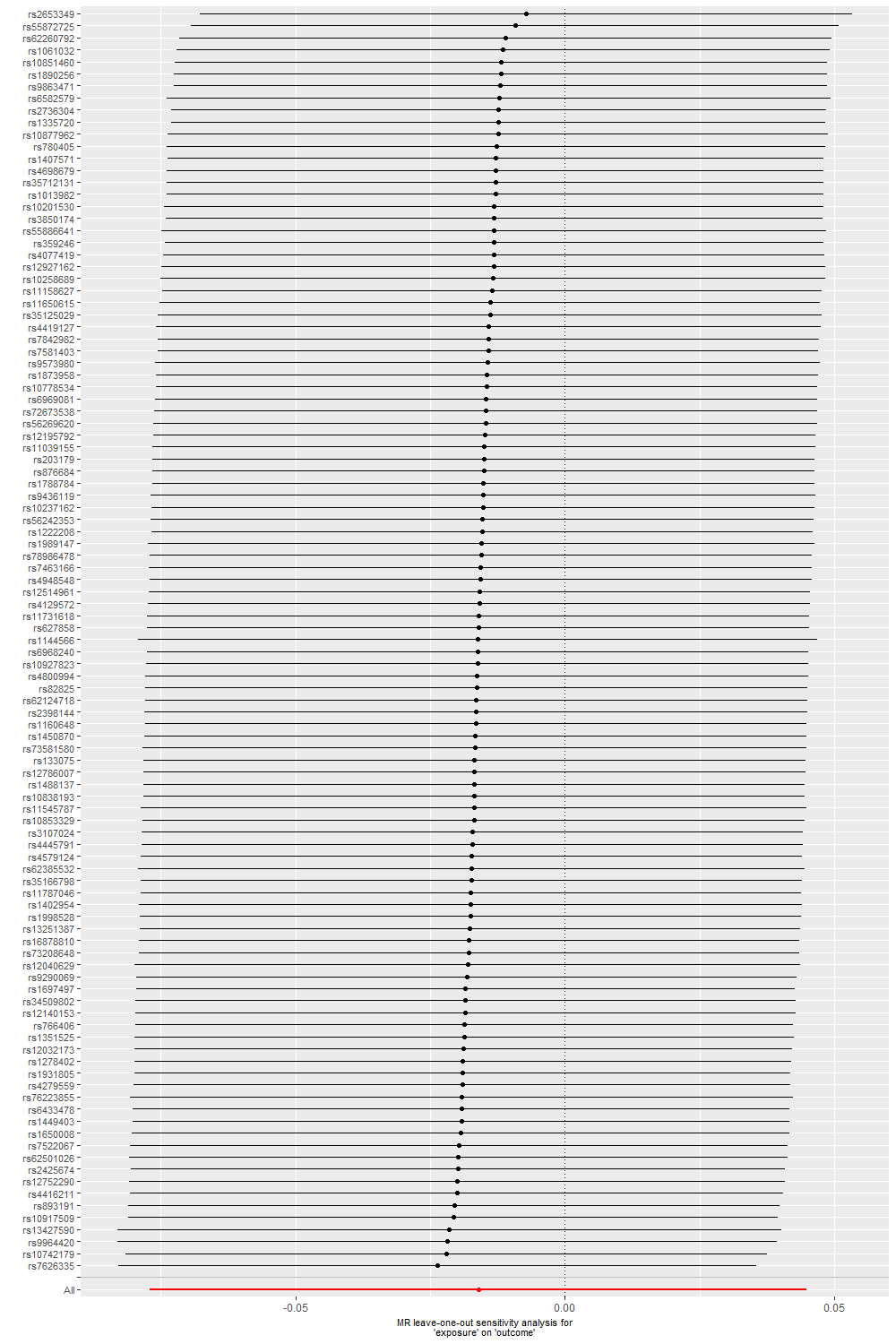

H. Macrosomia

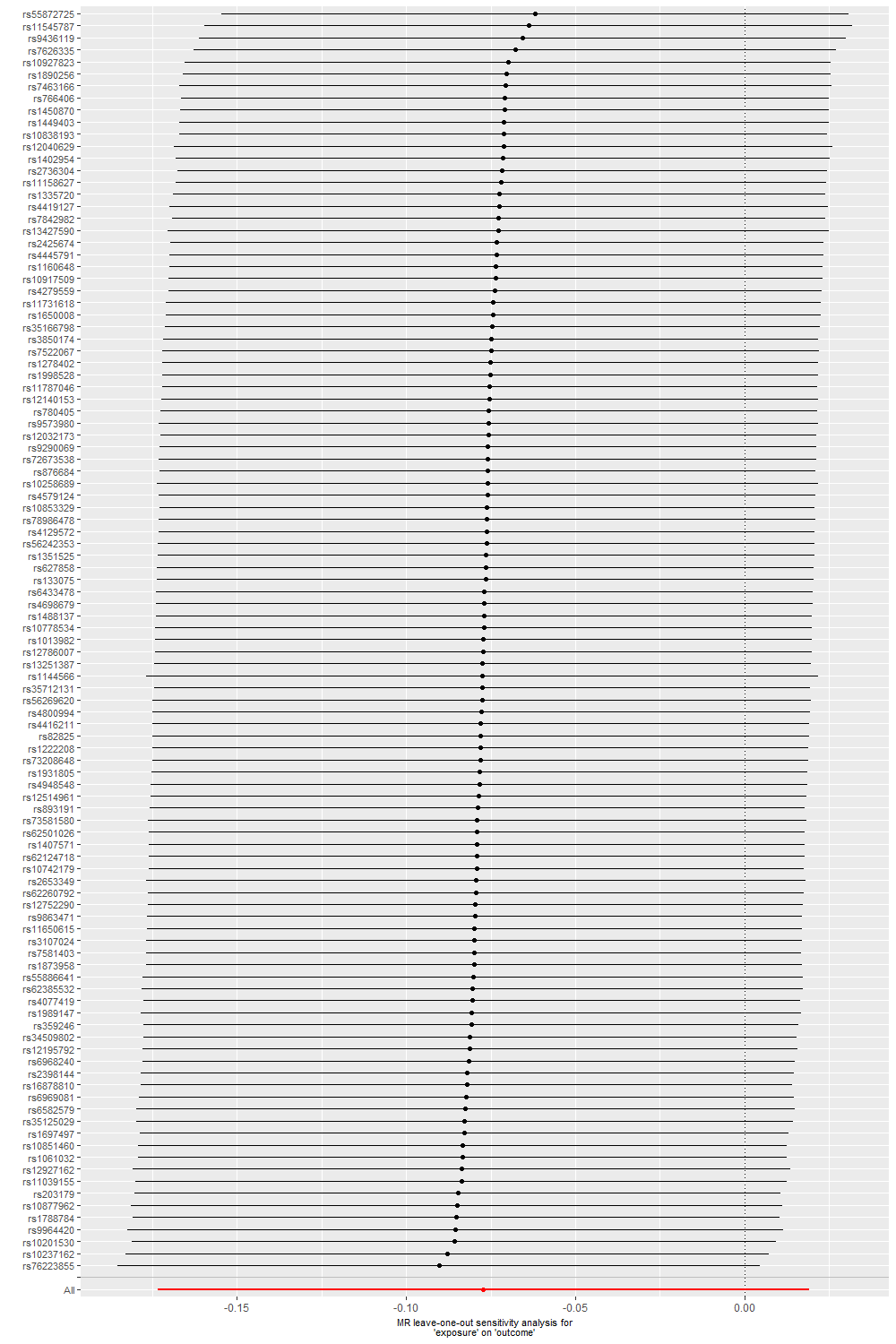

I. Offspring birthweight (grams)

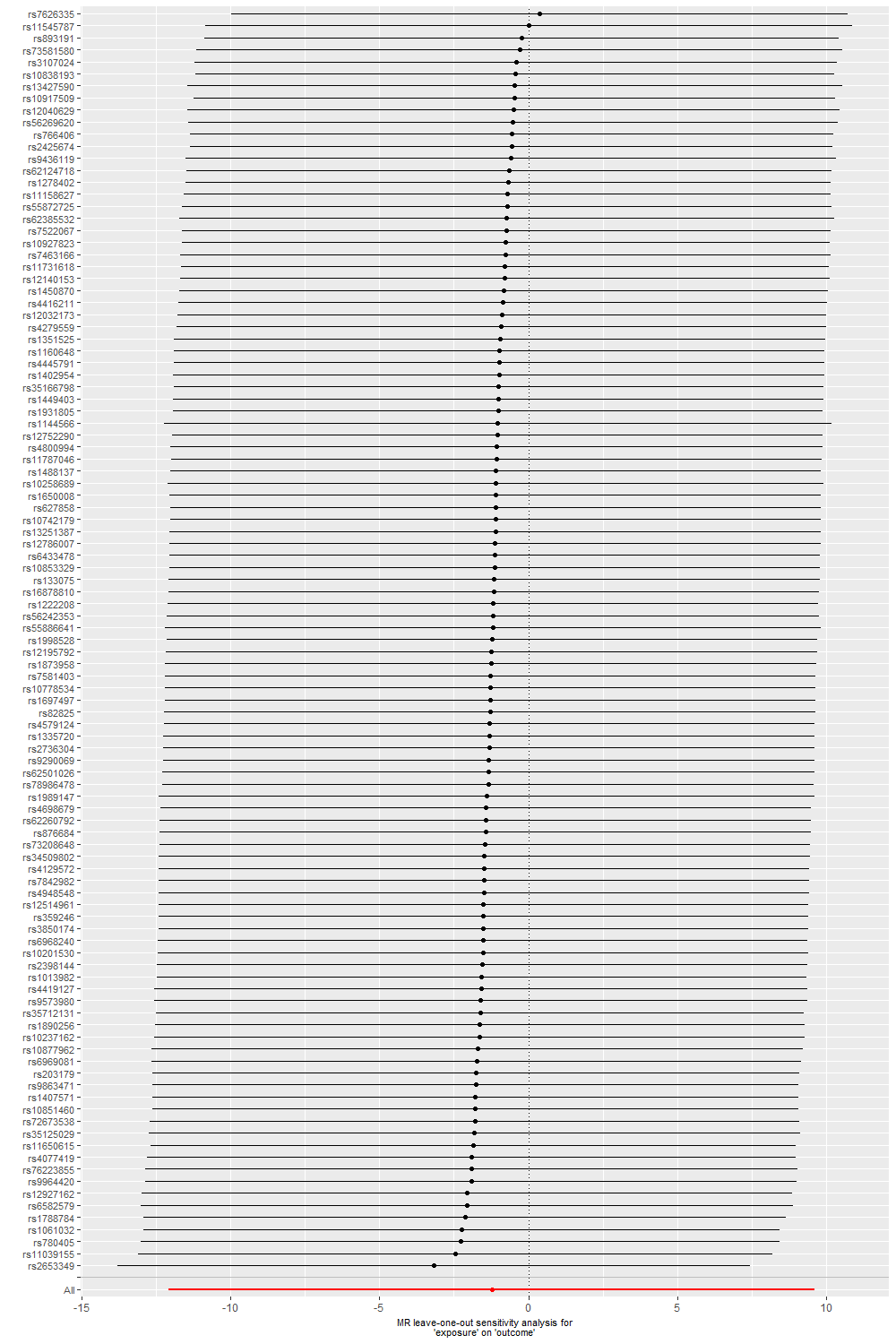

Abbreviations: ALSPAC, Avon Longitudinal Study of Parents and Children; BiB, Born in Bradford; MoBa, Norwegian Mother, Father and Child Cohort Study; MR, Mendelian randomization; SNP, single nucleotide polymorphisms; UKB, UK Biobank.

**eFigure 7. Comparisons of associations of 103 morning-preference SNPs with pregnancy and perinatal outcomes in the index pregnancy in Norwegian Mother and Child Cohort Study**

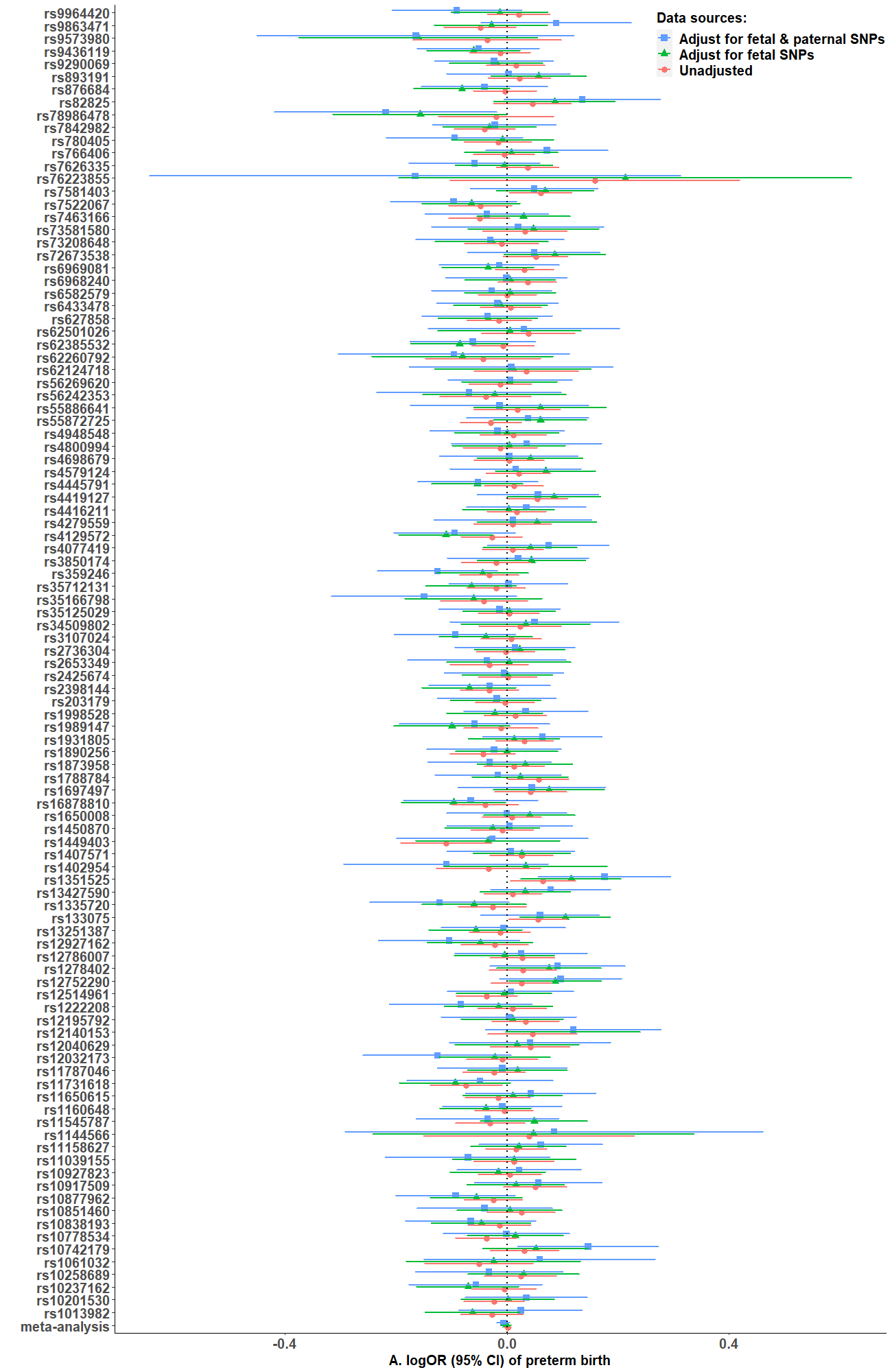

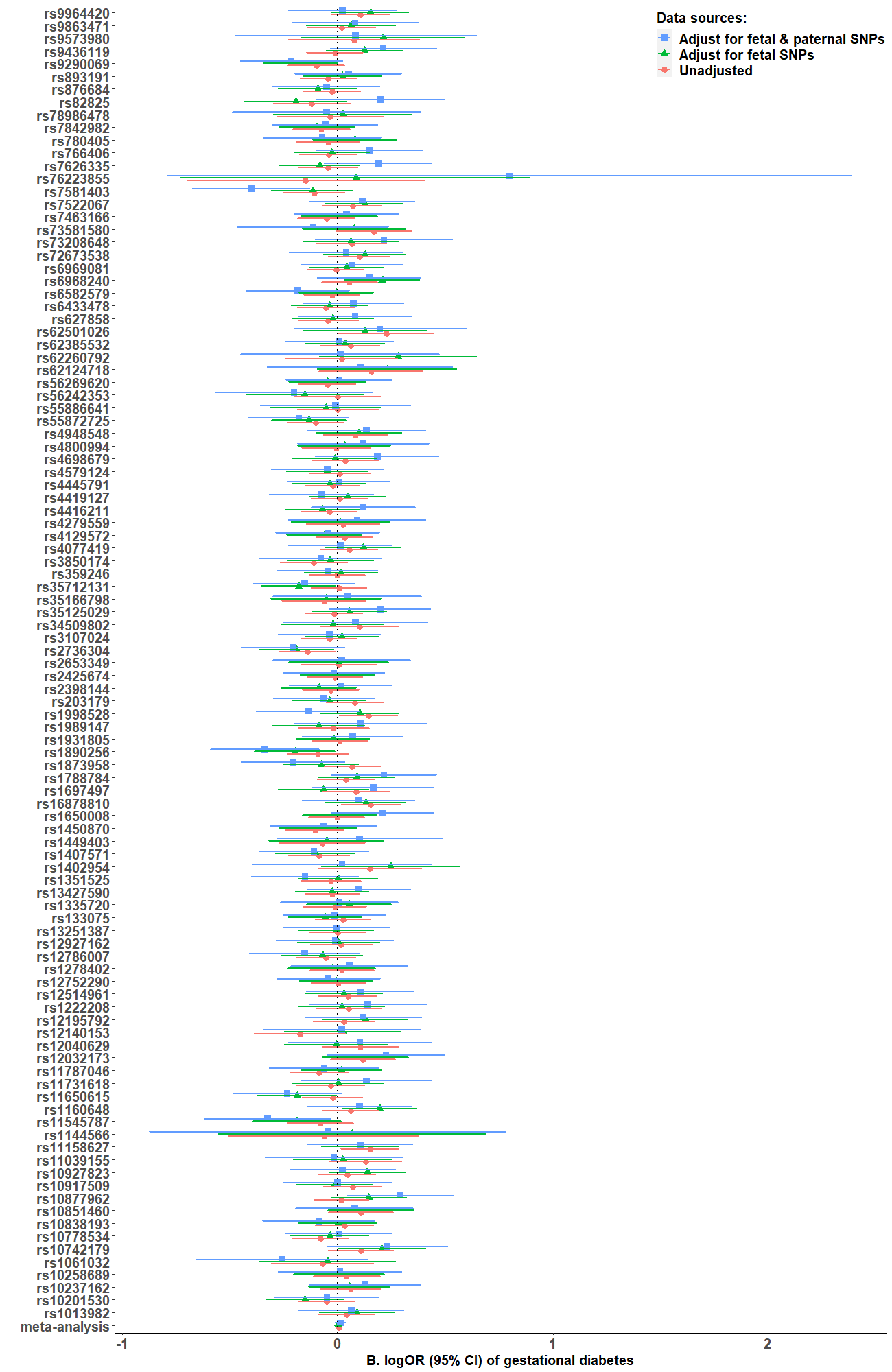

Statistical differences in SNPs (rs1873958 & rs1998528) – gestational diabetes associations were identified between adjustments for fetal & paternal SNPs versus no such adjustments (eTable 5 in Supplement).

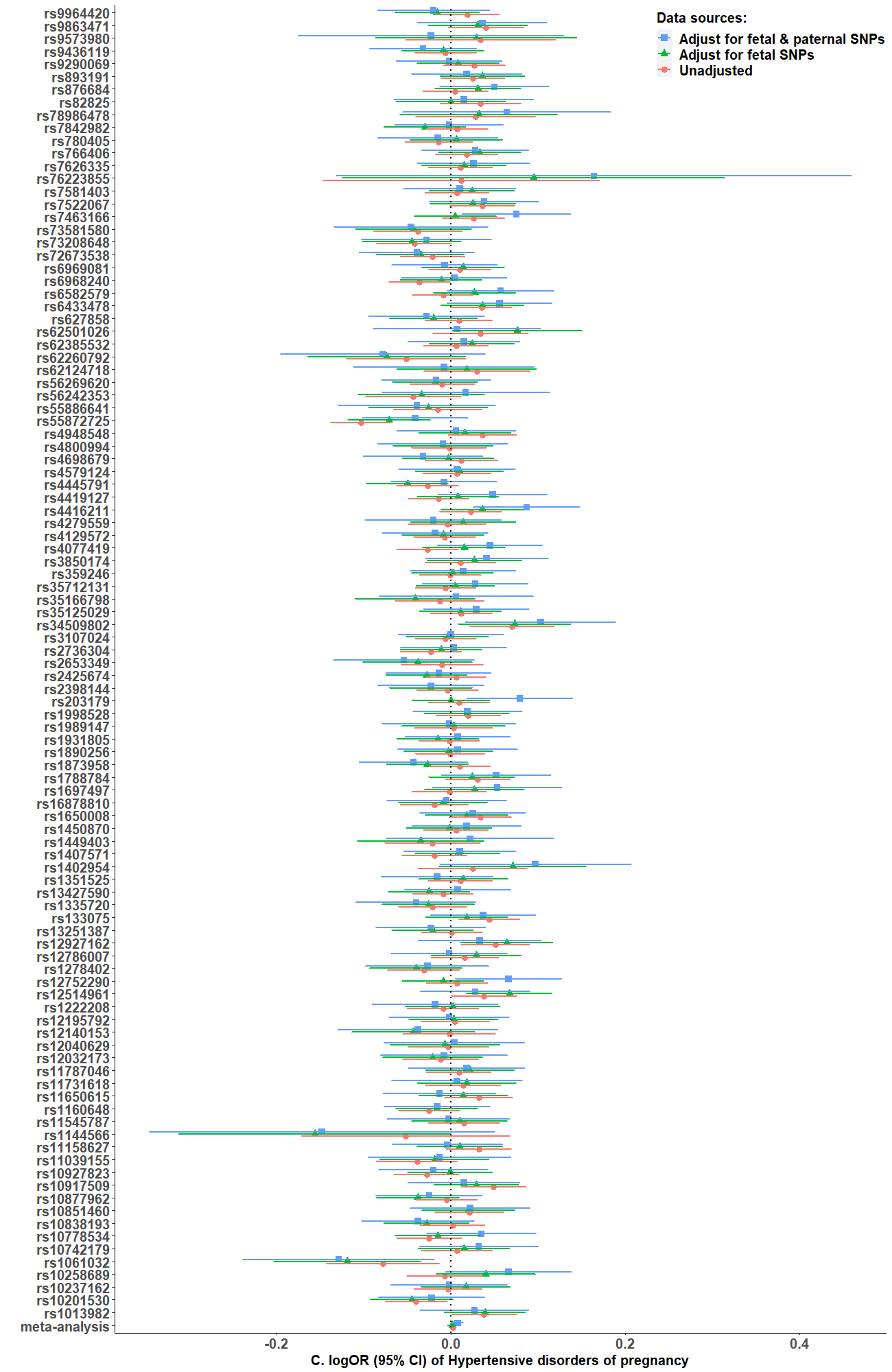

Statistical differences in rs4077419 – hypertensive disorders of pregnancy associations were identified between adjustments for fetal & paternal SNPs versus no such adjustments (eTable 5 in Supplement).

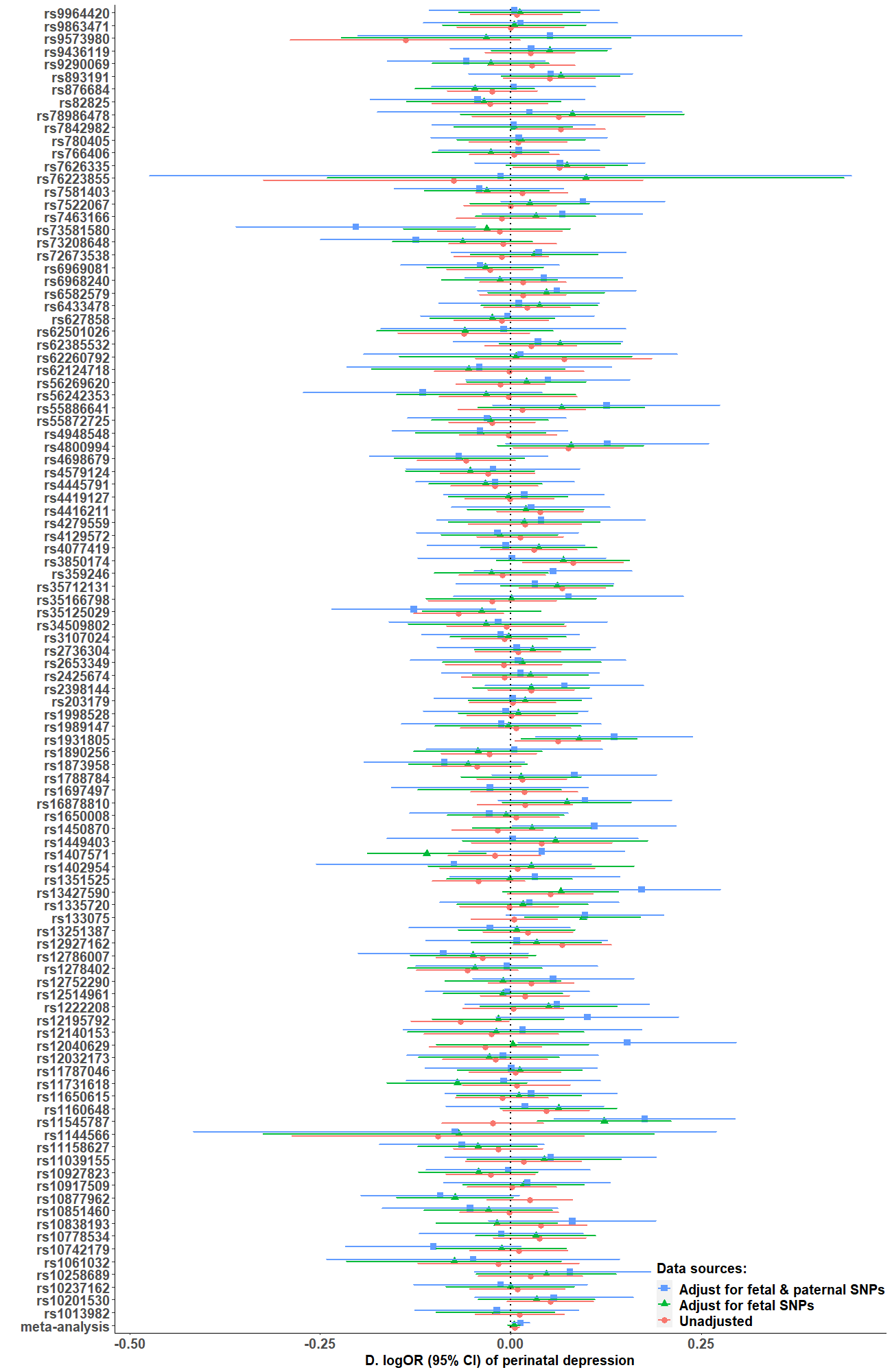

Statistical differences in SNPs – perinatal depression associations were identified between (1) adjustment for fetal SNP versus no such adjustment for rs10877962; and (2) adjustments for fetal & paternal SNPs versus no such adjustments for rs7358158 (eTable 5 in Supplement)..
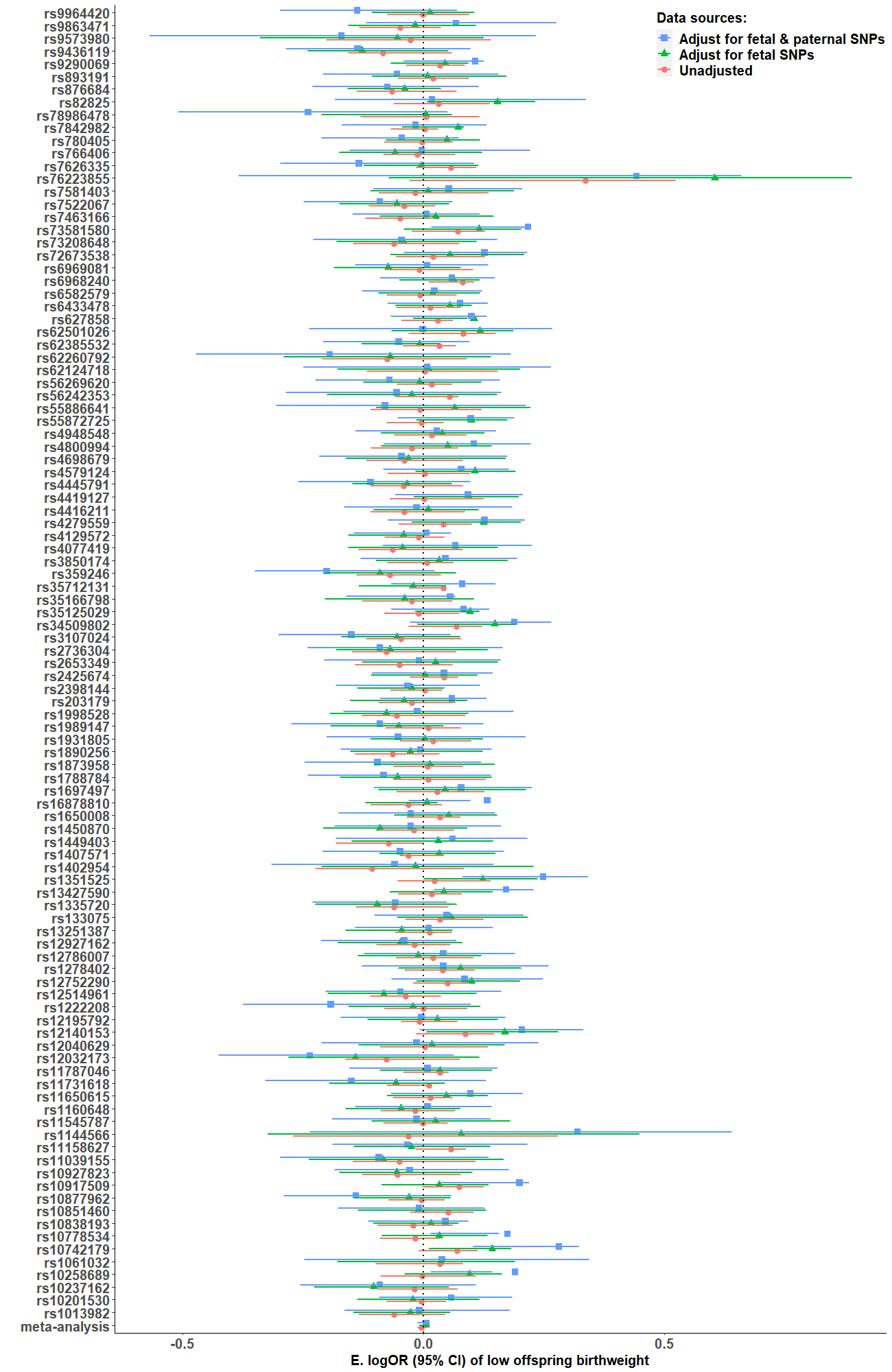

Statistical differences in rs7626335 – low offspring birthweight associations were identified between adjustments for fetal & paternal SNPs versus no such adjustments (eTable 5 in Supplement).

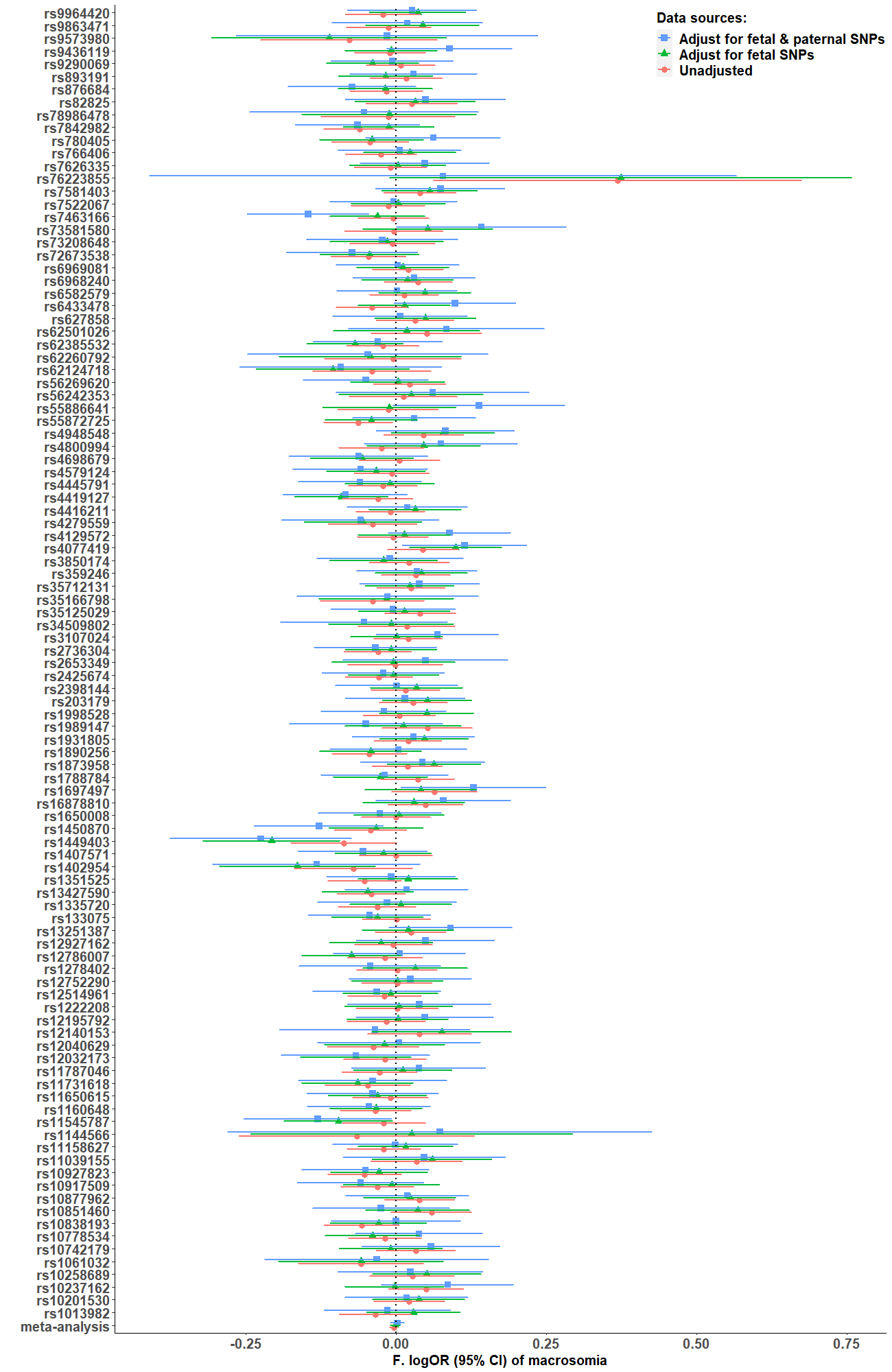

Statistical differences in SNPs (rs6433478 and rs7463166) – macrosomia associations were identified between adjustments for fetal & paternal SNPs versus no such adjustments (eTable 5 in Supplement).

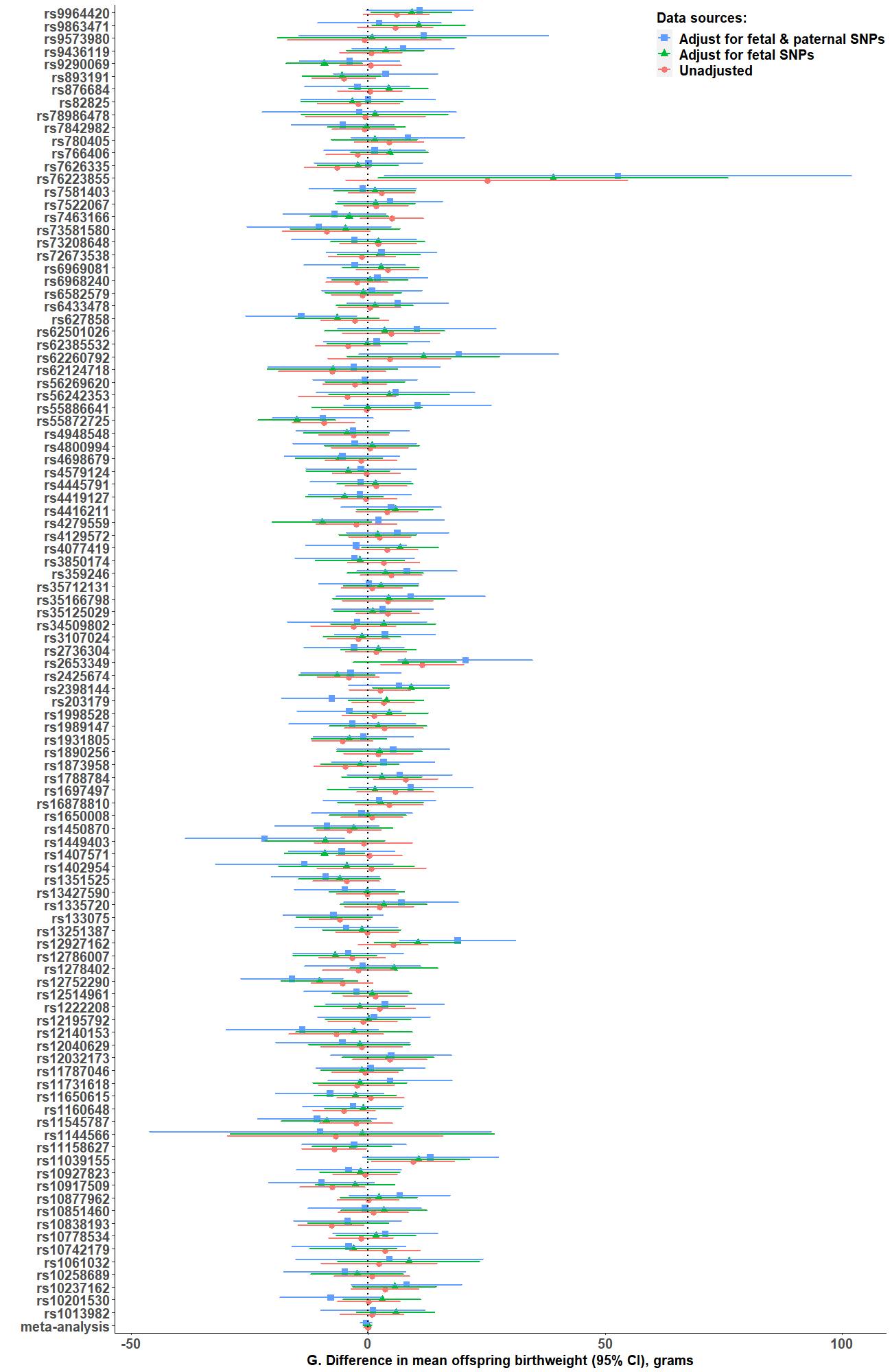

Statistical differences in rs1449403 – offspring birthweight associations were identified between adjustments for fetal & paternal SNPs versus no such adjustments (eTable 5 in Supplement).

Abbreviations: CI, confidence interval; OR, odds ratio; SNPs, single nucleotide polymorphisms.

**eFigure 8. Flow chart of each cohort**

A. ***UK Biobank (UKB)***

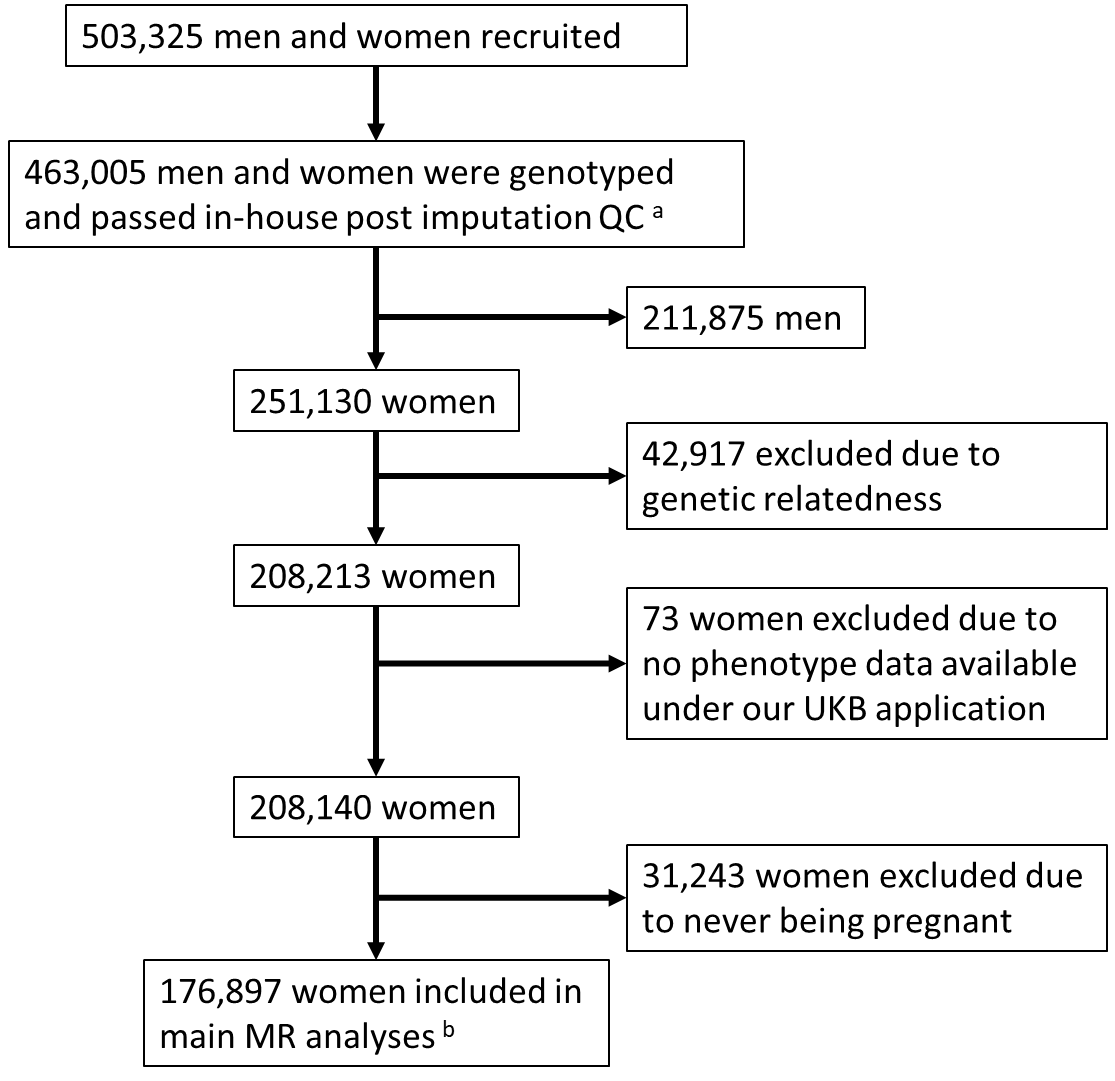

B. ***Avon Longitudinal Study of Parents and Children (ALSPAC)***

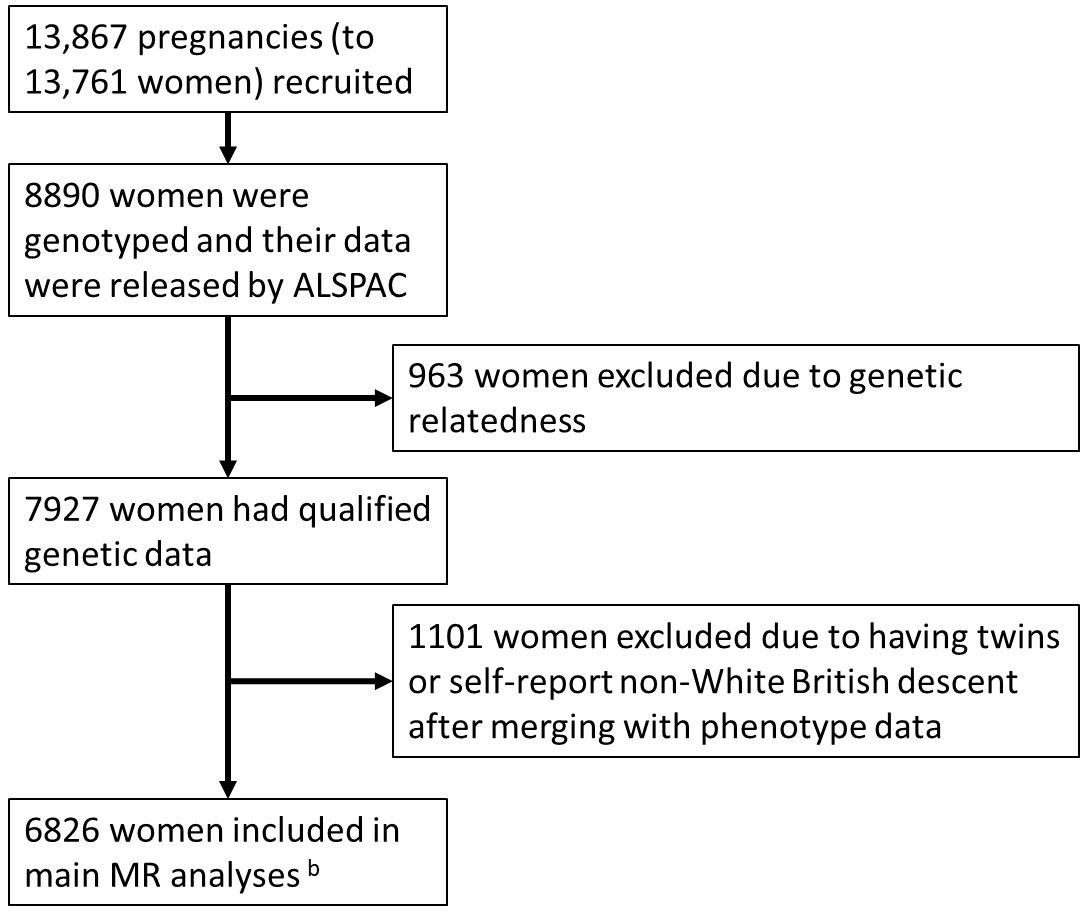

C. ***Born in Bradford (BiB)***

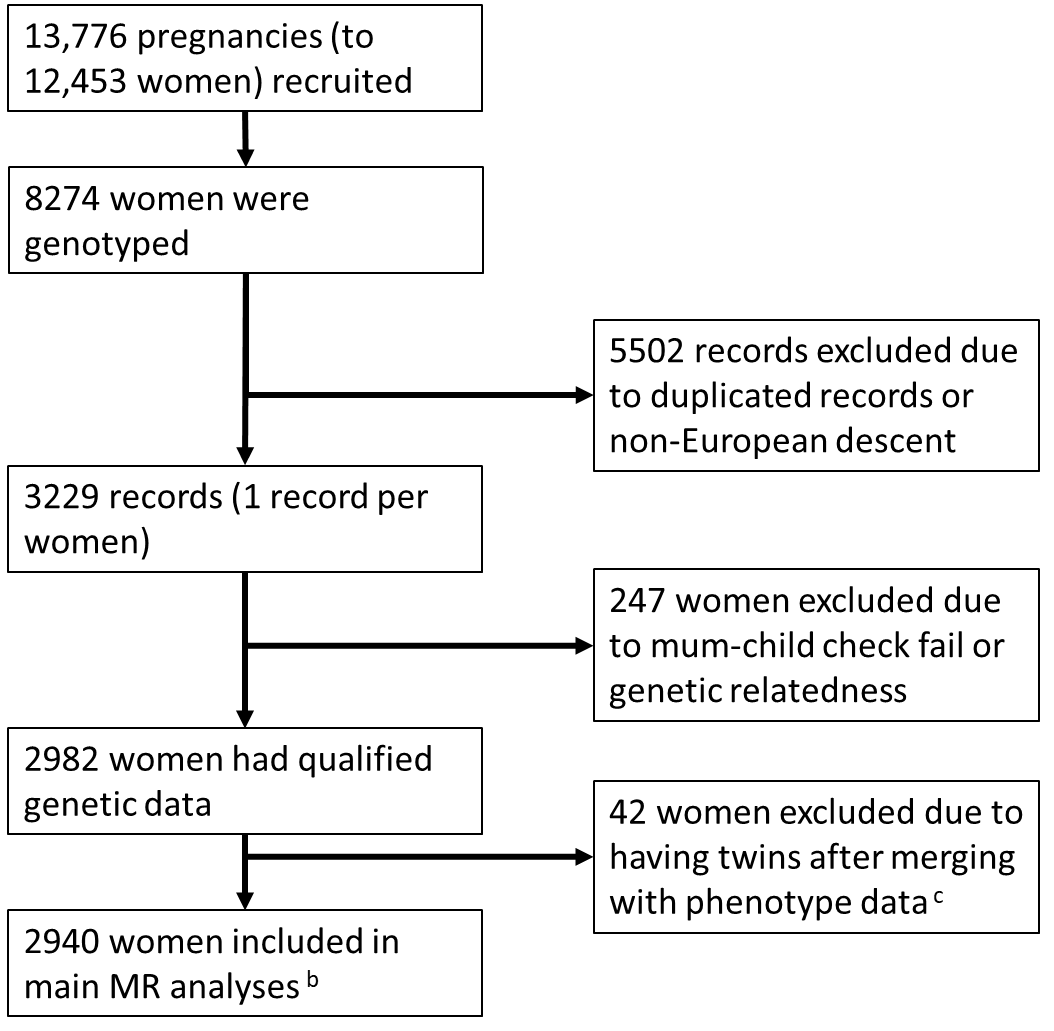

D. ***Norwegian Mother, Father and Child Cohort Study***

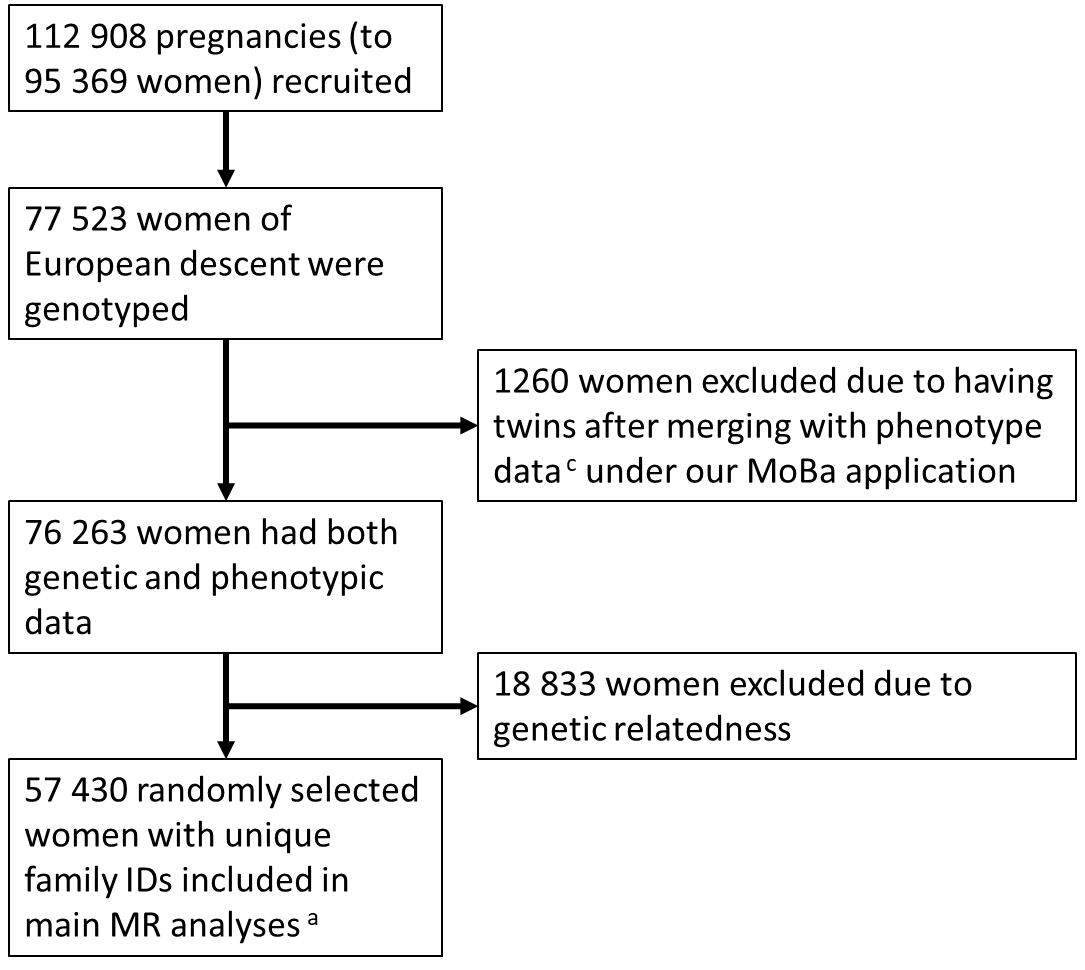

^a^ We used the imputed data released by UKB in March 2018, and applied in-house post-imputation QC ^7^.

^b^ The numbers of cases and controls for each pregnancy and perinatal outcomes are listed in Table 1.

^c^ We randomly selected one pregnancy per woman in the phenotype data if multiple pregnancies were recorded.

Abbreviation: MR, Mendelian randomization; QC, quality control.
